# From smartphone data to clinically relevant predictions: A systematic review of digital phenotyping methods in depression

**DOI:** 10.1101/2023.10.04.23296546

**Authors:** Imogen E. Leaning, Nessa Ikani, Hannah S. Savage, Alex Leow, Christian Beckmann, Henricus G. Ruhé, Andre F. Marquand

## Abstract

**Background:** Smartphone-based digital phenotyping enables potentially clinically relevant information to be collected as individuals go about their day. This could improve monitoring and interventions for people with Major Depressive Disorder (MDD). The aim of this systematic review was to investigate current digital phenotyping features and methods used in MDD.

**Methods:** We searched PubMed, PsycINFO, Embase, Scopus and Web of Science (10/11/2023) for articles including: (1) MDD population, (2) smartphone-based features, (3) validated ratings. Risk of bias was assessed using several sources. Studies were compared within analysis goals (correlating features with depression, predicting symptom severity, diagnosis, mood state/episode, other). Twenty-four studies (9801 participants) were included.

**Results:** Studies achieved moderate performance. Common themes included challenges from complex and missing data (leading to a risk of bias), and a lack of external validation.

**Discussion:** Studies made progress towards relating digital phenotypes to clinical variables, often focusing on time-averaged features. Methods investigating temporal dynamics more directly may be beneficial for patient monitoring.

European Research Council consolidator grant: 101001118, Prospero: CRD42022346264, Open Science Framework: https://osf.io/s7ay4

## Introduction

Major depressive disorder (MDD) is one of the most common and debilitating mental disorders worldwide, associated with a high personal and societal burden (Lim, et al., 2012). Moreover, MDD is often linked to a high recurrence risk (Buckman, et al., 2018), with over half of people who experience one depressive episode going on to have a subsequent episode (Burcusa & Iacono, 2007). Importantly, early signs of the development of symptoms or recurrence of depression are often not identified, which impedes timely preventive strategies. The broad integration of smartphones into people’s daily lives provides the unique opportunity to continuously and unobtrusively record behavioural dynamics in a naturalistic setting with high temporal resolution (Nelson & Allen, 2018). As such, it can offer insights into an individual’s mental state and could be useful for symptom monitoring and just-in-time preventive efforts in both non-clinical and clinical contexts (e.g., predicting symptom onset, or future recurrent episodes in patients with MDD).

Developing tools that leverage smartphone data to its full potential may therefore enable earlier identification and intervention before worsening of symptoms or recurrence of depression, leading ultimately to better outcomes. Smartphones can collect a wide range of behavioural information, for example geolocation data derived from the Global Positioning System (GPS), an individual’s use of social media or communication apps, general phone use/screen time, and typing-related data measuring psychomotor functioning or processing speed (Harari, Müller, Aung, & Rentfrow, 2017). Other technologies, such as wearable devices (e.g., wristbands), act as additional digital sources of behavioural or psychophysical measures. All these types of data can be used to create digital phenotypes, i.e., markers of behaviour or physiology calculated from digital measures, which could be indicative of clinically relevant behaviours. In this review we will focus on digital phenotypes for MDD created using smartphones, as these devices are now an integral and ubiquitous part of our daily lives. By installing monitoring tools on an individual’s own device, greater ecological validity may therefore be achieved than by using other devices as the risk of the monitoring altering participants’ behaviour may be lower than in studies where participants are required to adapt to wearing a device that they are not already accustomed to.

Digital phenotyping is a rapidly expanding technique (e.g., Farhan et al., 2016; Müller et al., 2021; Saeb et al., 2015; Ware et al., 2020), and a variety of different features have been explored in combination with various methods (e.g., logistic regression classifier, support vector machine, penalized logistic regression, random forest and XGBoost models) for classifying clinical labels or predicting clinically relevant information (e.g., depression scores). In order to understand how digital phenotyping can be used to better understand behavioural dynamics underlying MDD and to advance precision medicine endeavours aimed at earlier identification and/or intervention of (recurrent) depressive symptoms, high model performance is needed in addition to validation across multiple settings, including various symptom severities and lifestyles (e.g., working vs non-working populations). The general aim of this review was, therefore, to investigate the current state of digital phenotyping research for populations with MDD, in particular to establish what current methods are able to achieve in terms of their predictive power, and where subsequent efforts need to be focused to advance digital phenotyping in depression. Specifically, this systematic review aims to answer the following questions:

1. Of the different features that have been constructed from smartphone data, which are correlated with clinically relevant variables in the context of MDD?
2. What are the different methods that have been used for various depression prediction tasks using smartphone data and to what extent have these methods been successful?

First, we provide an overview and general evaluation of smartphone features (constructed across the included studies) that have been correlated with clinically relevant variables for depression (e.g., self-reported symptom scores), as feature construction is important for successful prediction models. Second, digital phenotyping studies in the field of MDD were compared that cover a variety of prediction tasks.

## Method

### Protocol and registration

This systematic review was guided by a protocol registered on Prospero (CRD42022346264) and the Open Science Framework (https://osf.io/s7ay4) and reported in line with the Preferred Reporting Items for Systematic Reviews and Meta-Analyses (PRISMA) guidelines.

### Information sources and search strategy

A comprehensive search was conducted on the following electronic databases: PubMed, PsycINFO, Embase, Scopus and Web of Science (November 10 2023). This search was restricted to studies published between January 2012 and November 2023 and included keywords related to (1) MDD, (2) digital phenotyping or monitoring, and (3) smartphones. For an overview of the exact keywords and search strings see S1 in supplementary materials.

### Selection of studies and eligibility criteria

Two authors (IL and NI) independently screened all titles and abstracts, and selective full texts screenings, to identify eligible papers for inclusion. Full texts of the selected papers were then examined to determine the final selection. In case of disagreement on inclusion, a third author (AM) was consulted to resolve divergent assessments. Central issues were discussed with all authors.

Studies were considered eligible if (1) passively collected smartphone data was utilised (e.g., GPS, use of communication apps, nearby Bluetooth devices) obtained with Android or iPhone smartphones, (2) passive data was collected over the course of the participants’ everyday lives (i.e., not collected during a laboratory session), and if (3) passively collected smartphone data was related to measures assessing depressive symptoms and/or diagnostic MDD status (e.g., self-reports, Ecological Momentary Assessments (EMA), structured clinical interviews) for the purpose of validating the digital phenotypes. Studies combining passively collected smartphone data with other data types, such as data from other wearable devices, were also included. Studies were excluded if (1) data was collected solely through means other than smartphones (e.g., wearable devices such as smart wristbands); (2) studies did not include participants with MDD (e.g., studies that included participants without a formal clinical diagnosis); (3) digital phenotyping-related studies with objectives not listed above (e.g., data collection verification studies); (4) reviews, overview articles, commentaries, etc.

### Data extraction, risk of bias assessment and quality assessment

Two authors (IL and NI) extracted data regarding study context, study sample, prediction goals, data acquisition, paradigms and analysis methods. For an overview of the exact data that was extracted see data extraction form S2 in supplementary materials.

Each included study was independently assessed by authors (IL and NI) for risk of bias using the criteria proposed by the Cochrane Collaboration Risk of Bias (RoB; Higgins, et al., 2016) and discrepancies were resolved with a third author (AM) (see Table S3 in supplementary materials). Five domains were rated as high risk, some concerns, low risk or unclear risk if there was risk of bias due to: (a) the used method for the randomisation sequence (selection bias); (b) allocation concealment (allocation bias); (c) blinding of participants and researchers (performance bias); (d) blinding of outcome assessment (detection bias); (e) incomplete outcome data (up to 10% drop out was rated as low risk) (attrition bias); (f) selective reporting (reporting bias). Items that were not relevant for a study were marked as ‘NA’.

Each included study was independently assessed by authors (IL and NI) for quality. Quality Assessment (QA) was assessed using items adapted from the guidelines created by Luo, et al. (2016), as well as from Benoit, Onyeaka, Keshavan, and Torous (2020) (see Table S4 in supplementary materials). The guidelines provided by Luo, et al. (2016) relate to machine learning methods, therefore, not all items were relevant/applicable for each included study. In these cases, relevant items were assessed and others listed as ‘NA’.

### Outcome measures

#### Outcomes included

1. Correlations between passively collected smartphone data and clinical measures.
2. Type of prediction strategy used to predict clinical labels or symptoms, and measures of performance of the prediction strategy. Studies were grouped by prediction goal (i.e., studies that predict depression symptom severity from smartphone features, clinical vs non-clinical labels or states (e.g., depressive state) and other analysis goals). Available metrics (e.g., classification accuracy, root mean squared error (RMSE)) were compared between studies against the backdrop of factors such as study population and included features, where informative, to assist meaningful comparisons.

In addition, participant (sample type) and study information relevant for RoB assessment and QA was extracted.

## Results

The search queries returned 24 eligible studies with several analysis and prediction goals (see Figure 1 for PRISMA flow chart. Examples of exclusion based on study design included studies that utilised solely digital phenotypes calculated from digital devices other than smartphones, or that used digital phenotypes calculated from actively collected smartphone data. Examples of exclusion based on publication type included published protocols, reviews and dissertations). Characteristics of included studies are provided in Table 1 and general methodological information for these studies is summarised in Table 2. Specific methodological information per analysis goal is summarised in Tables 3-6. Included studies used features calculated from a range of sensor streams, for example count-based and statistical features reflecting mobility sensors (e.g., GPS data) and communication sensors (e.g., app-based data). Studies were grouped by analysis goal to allow for comparison of methods with similar objectives. To this end, to investigate our first research question we first compared studies that correlated individual passive smartphone features with depression symptom severity (Cao, et al., 2020; Sun, et al., 2023; Sverdlov, et al., 2021; Wasserzug, et al., 2023; Zhang, et al., 2022; Zou, et al., 2023). To address our second research question, we then shifted our focus to the different methods that have been used for various depression prediction tasks, investigating the methods used for predicting symptom severity (Braund, et al., 2022; Cao, et al., 2020; Faurholt-Jepsen, et al., 2022; Kathan, et al., 2022; Pedrelli, et al., 2020; Pellegrini, et al., 2022; Sverdlov, et al., 2021; Zhang, et al., 2021; Zhang, et al., 2022). Somewhat unexpectedly, two studies aimed to predict specific smartphone features from ratings of depression (Laiou, et al., 2022; Tønning, Faurholt-Jepsen, Frost, Bardram, & Kessing, 2021). We then shifted our focus towards studies that aimed to classify participants into different diagnostic classes and mood states (Bai, et al., 2021; Cho, et al., 2019; Faurholt-Jepsen, et al., 2022; Lee, et al., 2023; Kim, et al., 2023; Sverdlov, et al., 2021; Wasserzug, et al., 2023). Some studies contained unique goals that were not shared with the other studies, and these goals are considered in a separate section. We also compared some key methodological choices, such as feature selection and processing, dimension reduction, and handling of missing data.

**Figure 1.**
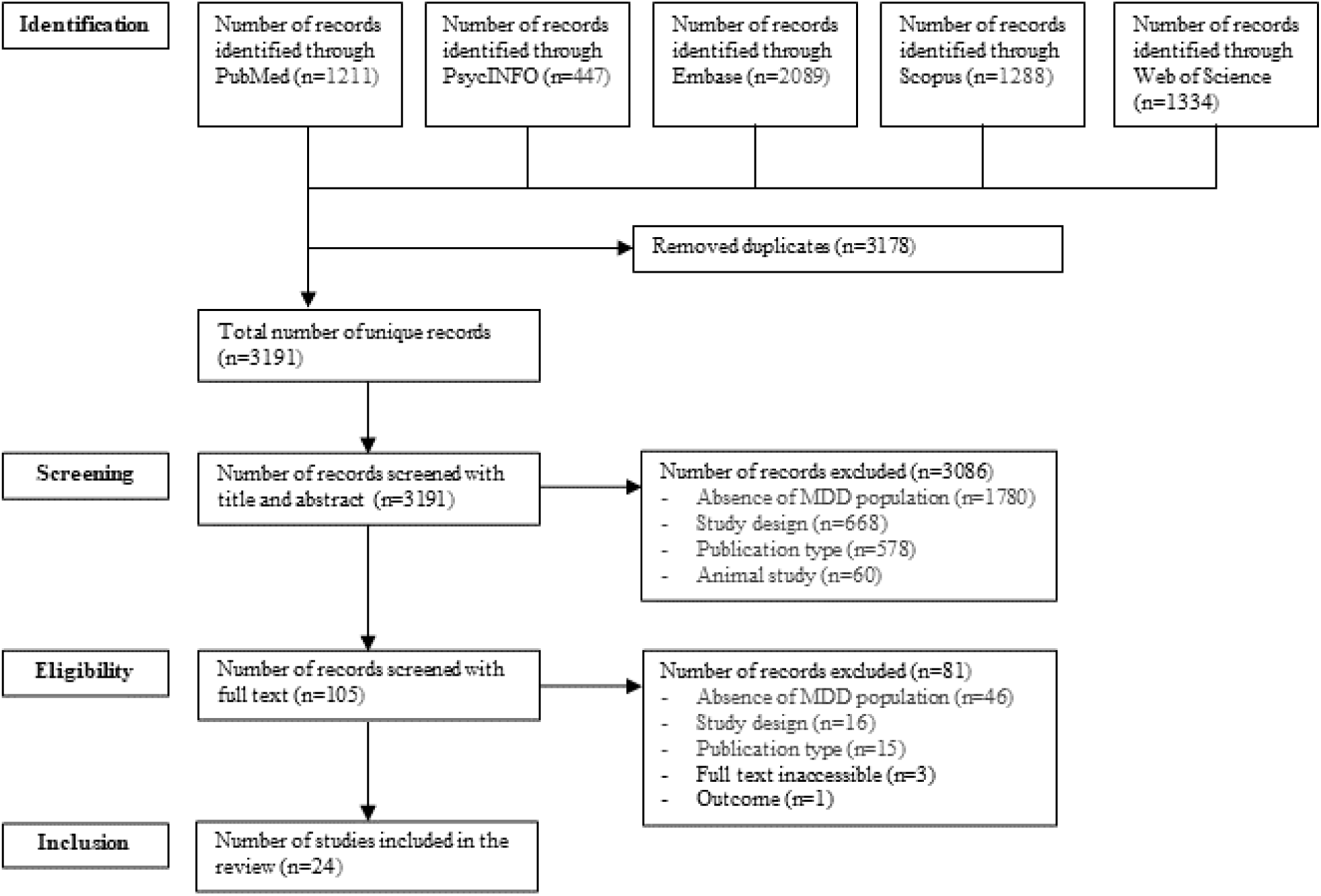
Flowchart of selection and inclusion process following the PRISMA Statement.

**Table 1.** Characteristics of Included Studies.

| Author (year of publication) | Country | Population | % Female | Age (*M*) | Ethnicity | Sample size |
| --- | --- | --- | --- | --- | --- | --- |
| Bai et al. (2021) | China | MDD | NP | NP | NP | 334 |
| Braund et al. (2022) | Australia | MDD (n = 79); BD (n = 42) | 65.3 | 41.4 | NP | 121 |
| Cao et al. (2020) | USA | MDD | 84.6 | 14.93 | NP | 13 |
| Cho et al. (2019) | Korea | MDD (n = 18); BD I (n = 18); BD II (n = 19) | 49.1 | 25.92 | NP | 55 |
| Emden et al. (2020) | Germany | MDD (n = 409); BD (n = 48); AD (n = 58); PD (n = 21); HC (n = 458) | 67.3 | 35.99 | NP | 997 |
| Faurholt-Jepsen et al. (2022) | Denmark | MDD (n = 75); BD (n = 65) | 56.6 | 44.2 | NP | 140 |
| Fujino et al. (2023) | Japan | MDD | 30.5% | 41.2 | NP | 2143 |
| Kathan et al. (2022) | Germany | MDD | 81.2% | 32.8 | NP | 16 |
| Kim et al. (2023) | Republic of Korea | MDD (n = 24); HC (n = 10) | 64.7% | 14.9 | NP | 34 |
| Knights et al. (2023) | USA | MDD (n = 955); BD (n = 471); SZ (n = 152); personality disorder (n = 81); other (n = 685); missing (n = 8) | 73.6% | 56.7 | White = 48.9%; African American = 10.5%; Hispanic/Latino = 3.4%; multiracial = 2.1%; did not specify = 34.3% | 2352 |
| Laiou et al. (2021) | UK, the Netherlands, Spain | MDD | 75.0 | 48 (median) | NP | 164 |
| Lee et al. (2023) | Republic of Korea | MDD (n = 95); BD I (n = 78); BD II (n = 97) | 54.4% | 23.3 | NP | 270 |
| Matcham et al. (2022) | UK, the Netherlands, Spain | MDD (n = 378); HC (n = 245) | 75.6 | 46.4 | NP | 623 |
| Pedrelli et al. (2020) | USA | MDD | 74.0 | 33.7 | White = 71%; Hispanic/Latino = 23%; Asian = 16%; Haitian/Black/African-American = 12%; American Indian/Alaskan = 3%; mixed-race = 6%; other = 3% | 41 |
| Pellegrini et al. (2021) | USA | MDD (n = 10); BD (n = 10); SZ (n = 10); HC (n = 11) | 63.0 | 43.0 | White = 71%; African-American = 20%; Asian = 7%; Other = 2% | 41 |
| Siddi et al. (2022) | UK, Spain, the Netherlands | MDD | 74.51% | 47.73 | NP | 255 |
| Sun et al. (2023) | UK, Spain, the Netherlands. | MDD | 74.5% | 50.0 (median) | NP | 479 |
| Sverdlov et al. (2021) | the Netherlands | MDD (n = 20); HC (n = 20) | 33.0 | 31.2 | White = 87.5%; Mixed = 7.5%; Asian = 2.5%; Black or African-American = 2.5% | 40 |
| Tønning et al. (2021) | Denmark | MDD | 52.7 | 44.4 | NP | 74 |
| Wasserzug et al. (2023) | Israel | MDD (n = 40); HC (n = 104) | 50.7% | NP | NP | 144 |
| Zhang et al. (2021) | Spain, the Netherlands, UK | MDD | 74.1 | 51.0 (median) | NP | 316 |
| Zhang et al. (2022) | Spain, the Netherlands, UK | MDD | 74.1 | 50.0 (median) | NP | 290 |
| Zhang et al. (2023) | Spain, the Netherlands, UK | MDD | 75.7% | 49 (median) | KCL: white = 84.3%; VUMC: white = 92.4%; CIBER: NP | 614 |
| Zou et al. (2023) | China | MDD | 65.7% | 31.6 | NP | 245 |

AD = Anxiety Disorder; BD (I or II) = Bipolar Disorder (Type 1 or Type 2); HC = Healthy Control; M = Mean; MDD = Major Depressive Disorder; PD = Psychotic Disorder; SZ = Schizophrenia/Schizoaffective Disorder; NP = Not Provided

**Table 2.**
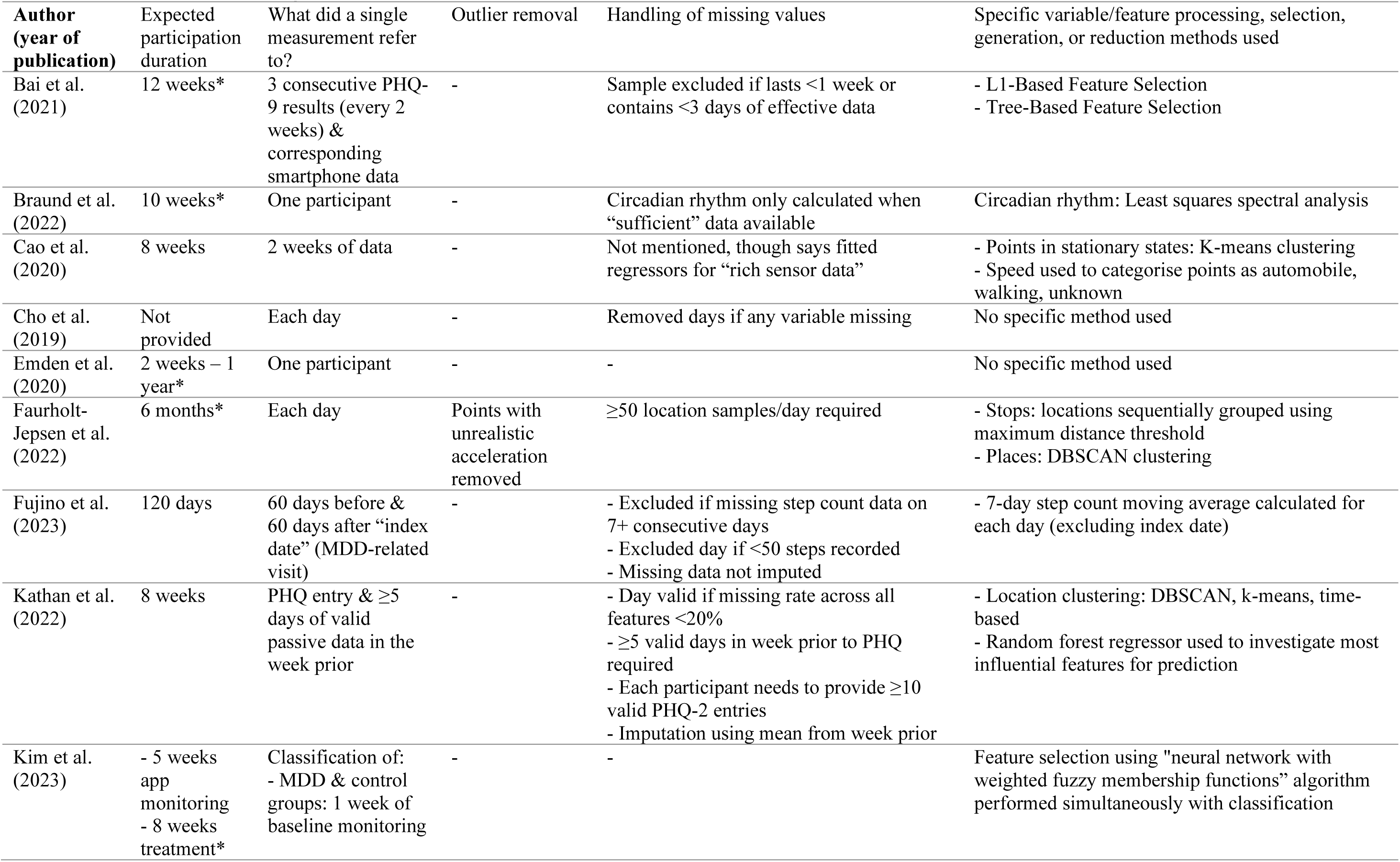
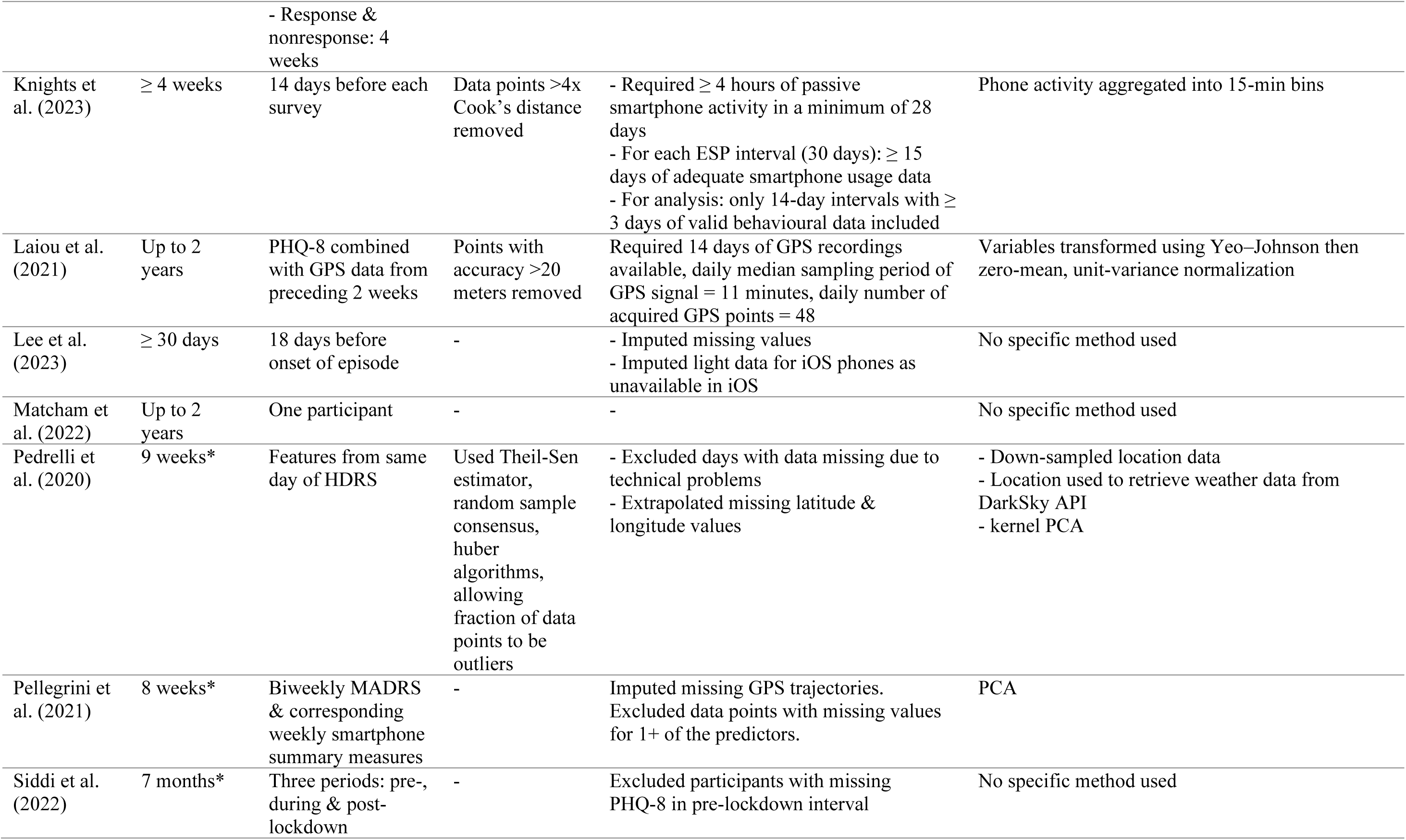
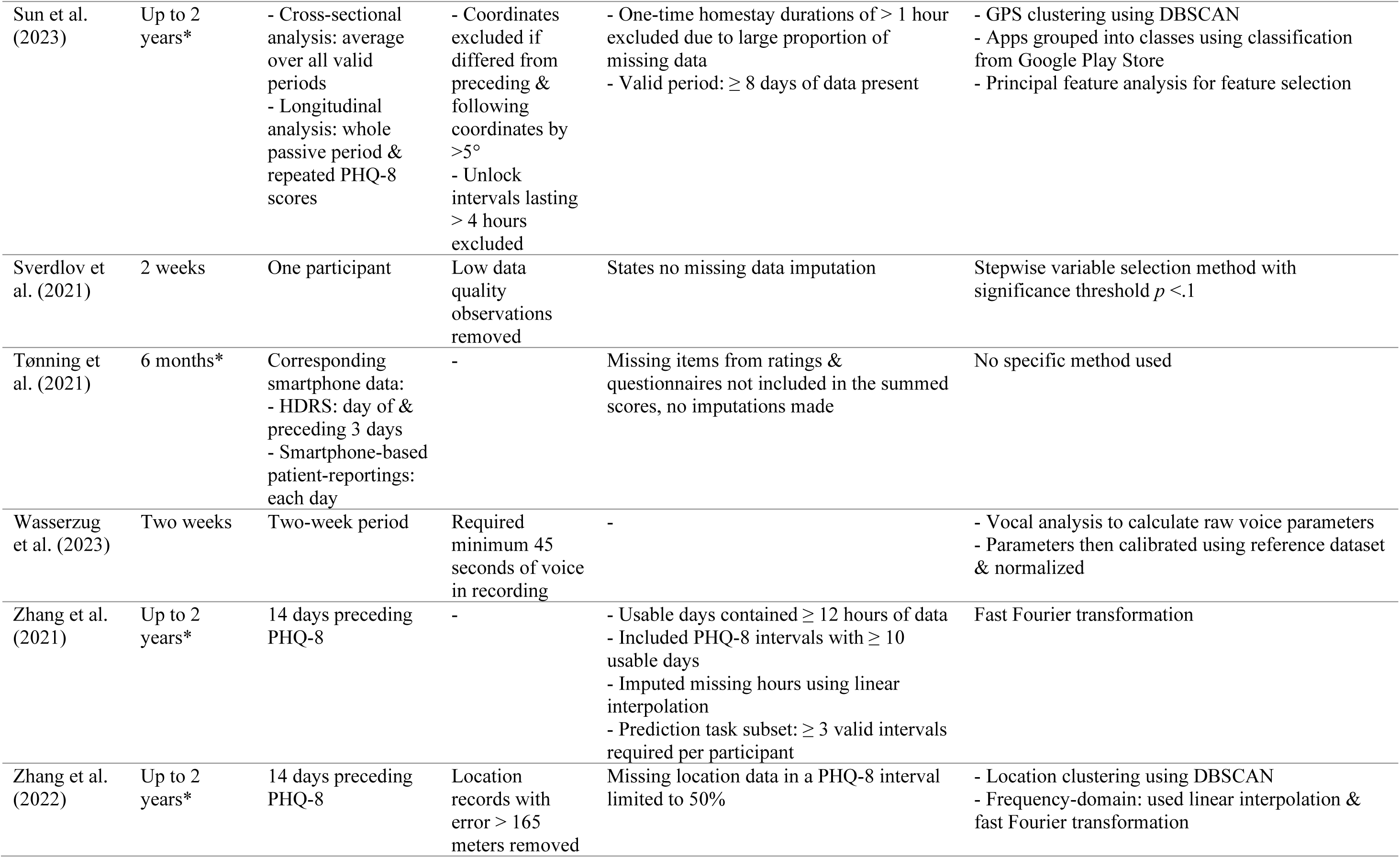
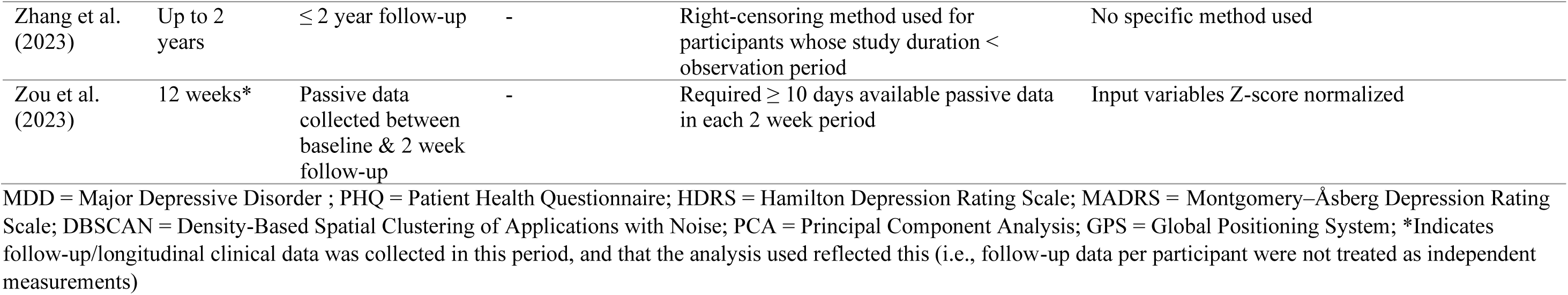
General Methodological Information Of All Included Studies.

**Table 3.**
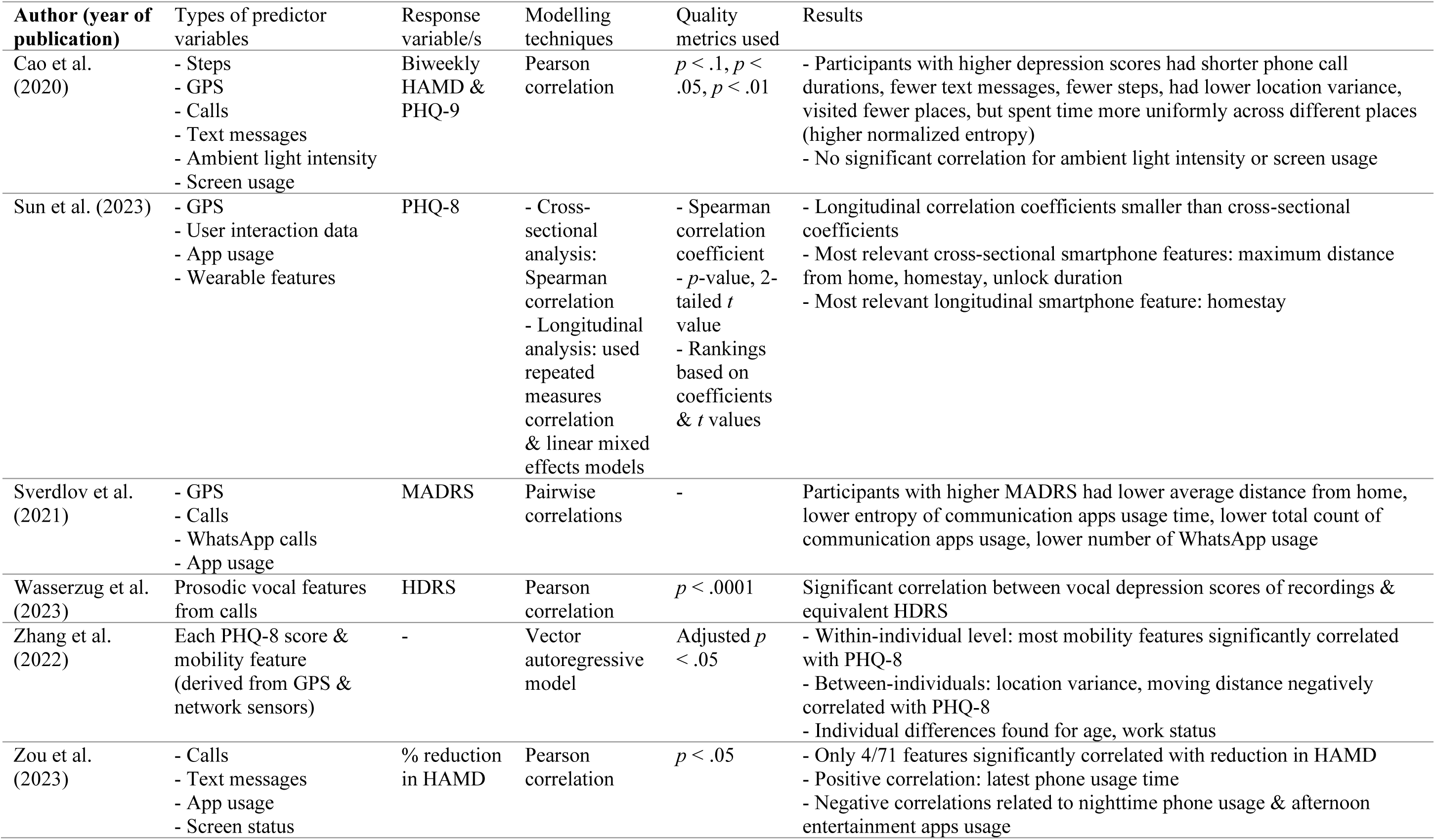
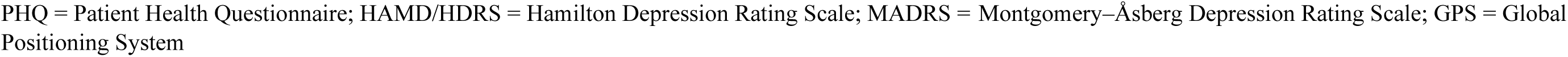
Methodological Details Of Studies Correlating Passive Smartphone Features With Depression Symptom Severity.

| Author (year of publication) | Types of predictor variables | Response variable/s | Modelling techniques | Quality metrics used | Results |
| --- | --- | --- | --- | --- | --- |
| Cao et al. (2020) | - Steps<br>- GPS<br>- Calls<br>- Text messages<br>- Ambient light intensity<br>- Screen usage | Biweekly HAMD & PHQ-9 | Pearson correlation | $p < .1, p < .05, p < .01$ | - Participants with higher depression scores had shorter phone call durations, fewer text messages, fewer steps, had lower location variance, visited fewer places, but spent time more uniformly across different places (higher normalized entropy)<br>- No significant correlation for ambient light intensity or screen usage |
| Sun et al. (2023) | - GPS<br>- User interaction data<br>- App usage<br>- Wearable features | PHQ-8 | - Cross-sectional analysis: Spearman correlation<br>- Longitudinal analysis: used repeated measures correlation & linear mixed effects models | - Spearman correlation coefficient<br>- $p$-value, 2-tailed $t$ value<br>- Rankings based on coefficients & $t$ values | - Longitudinal correlation coefficients smaller than cross-sectional coefficients<br>- Most relevant cross-sectional smartphone features: maximum distance from home, homestay, unlock duration<br>- Most relevant longitudinal smartphone feature: homestay |
| Sverdlov et al. (2021) | - GPS<br>- Calls<br>- WhatsApp calls<br>- App usage | MADRS | Pairwise correlations | - | Participants with higher MADRS had lower average distance from home, lower entropy of communication apps usage time, lower total count of communication apps usage, lower number of WhatsApp usage |
| Wasserzug et al. (2023) | Prosodic vocal features from calls | HDRS | Pearson correlation | $p < .0001$ | Significant correlation between vocal depression scores of recordings & equivalent HDRS |
| Zhang et al. (2022) | Each PHQ-8 score & mobility feature (derived from GPS & network sensors) | - | Vector autoregressive model | Adjusted $p < .05$ | - Within-individual level: most mobility features significantly correlated with PHQ-8<br>- Between-individuals: location variance, moving distance negatively correlated with PHQ-8<br>- Individual differences found for age, work status |
| Zou et al. (2023) | - Calls<br>- Text messages<br>- App usage<br>- Screen status | % reduction in HAMD | Pearson correlation | $p < .05$ | - Only 4/71 features significantly correlated with reduction in HAMD<br>- Positive correlation: latest phone usage time<br>- Negative correlations related to nighttime phone usage & afternoon entertainment apps usage |

PHQ = Patient Health Questionnaire; HAMD/HDRS = Hamilton Depression Rating Scale; MADRS = Montgomery–Åsberg Depression Rating Scale; GPS = Global Positioning System

### Correlation between passive smartphone features and depressive symptom severity

Six studies were identified that investigated correlations between features derived from passively-collected smartphone data (e.g., total amount of time spent at home, number of unique phone call partners/day, total number and duration of phone calls) and depressive symptom severity quantified using sum scores of self-reports, such as the Patient Health Questionnaire (PHQ), and clinician-rated Montgomery–Åsberg Depression Rating Scale (MADRS) and Hamilton Depression Rating Scale (HDRS). These studies are summarised in Table 3. Sverdlov, et al. (2021) and Cao, et al. (2020) investigated various features related to participants’ communication behaviours. Sverdlov, et al. (2021) found that more severe depressive symptom scores tended to have lower entropy of usage time of communication apps, lower total count of communication apps usage, and lower WhatsApp usage. Cao, et al. (2020) found that a higher depression score is significantly correlated with lower social interaction levels (i.e., shorter phone call durations, fewer text messages sent), which seems consistent with Sverdlov, et al.’s (2021) findings from app-based investigations, despite the differences in data type. Wasserzug, et al. (2023) also found a significant correlation between vocal depression scores derived from prosodic vocal features and depressive symptom scores.

In terms of general phone usage, Sun, et al. (2023) found that unlock duration was positively correlated with depression score, and Zou, et al. (2023) found a positive correlation between latest phone usage time and depression score, and a negative correlation with night time phone usage and with afternoon entertainment app usage. However, no significant correlation between symptom scores and smartphone screen usage was found by Cao, et al., 2020, and Zou, et al., 2023 noted that only a small number of features were found to have a significant correlation.

Various mobility features have also been related to individuals’ symptom scores. Sverdlov, et al. (2021) found that participants with higher symptom scores tended to maintain a lower average distance from home than participants with less severe symptom scores. Similarly, Sun, et al. (2023) found a negative correlation between maximum distance travelled from home and depression symptom scores, and a positive correlation with homestay duration. Cao, et al. (2020) also found that individuals with higher symptom scores demonstrated lower mobility, as indicated by decreased step count, fewer places visited, and lower location variance, spending their time more uniformly across different places. Zhang, et al. (2022) investigated correlations between mobility features and symptom scores provided by their vector autoregressive model. They found that within individuals, the proportion of time spent at their home location/s (‘homestay’) and short-term rhythm (i.e., behavioural rhythms with frequency higher than one day, e.g., for many people, going to and from their home) were positively correlated with symptom scores. Other features, for example long-term rhythm (behavioural rhythms with frequency less than one day, e.g., a weekly grocery shop) and circadian rhythm, were negatively correlated with symptom scores. Between individuals, only location variance and moving distance were negatively correlated with symptom scores. Overall, studies generally identified that higher symptom scores were associated with lower mobility.

Regarding the significance of the results and correction for multiple testing, a range of *p*-value significance thresholds were used in the studies, and it was mostly unclear if thresholds were corrected for multiple testing (see Table 3). Overall, correlations were generally weak to moderate, with Cao, et al. (2020) reporting the strongest correlation magnitude of approximately 0.65 for daily step count.

### Predicting depression symptom severity

Several studies investigated the possibility of using passively-collected smartphone data to predict depression symptom severity (displayed in Table 4; symptom scale ranges are provided in Table S5 in supplementary materials to assist interpretation of model performance). Linear regression and linear mixed-effect regression models were popular choices for this prediction goal. Sverdlov, et al. (2021) used a subset of the communication and mobility features discussed in the preceding section to predict MADRS scores using a multiple linear regression model. The correlation between observed and predicted scores calculated in leave-one-out cross-validation was r = 0.43, showing a moderate correlation. Pellegrini, et al. (2022) conducted a Principal Component Analysis on a set of GPS and accelerometer features, and used the first principal component as a predictor in their linear mixed models. Pellegrini, et al. (2022) investigated various models with and without this passive smartphone feature and a baseline depressive symptom score, demonstrating that including a smartphone feature did not improve the prediction of MADRS scores, but instead was comparable to predictions by models using only questionnaire data (RMSE = 4.30, 4.27 respectively).

**Table 4.**
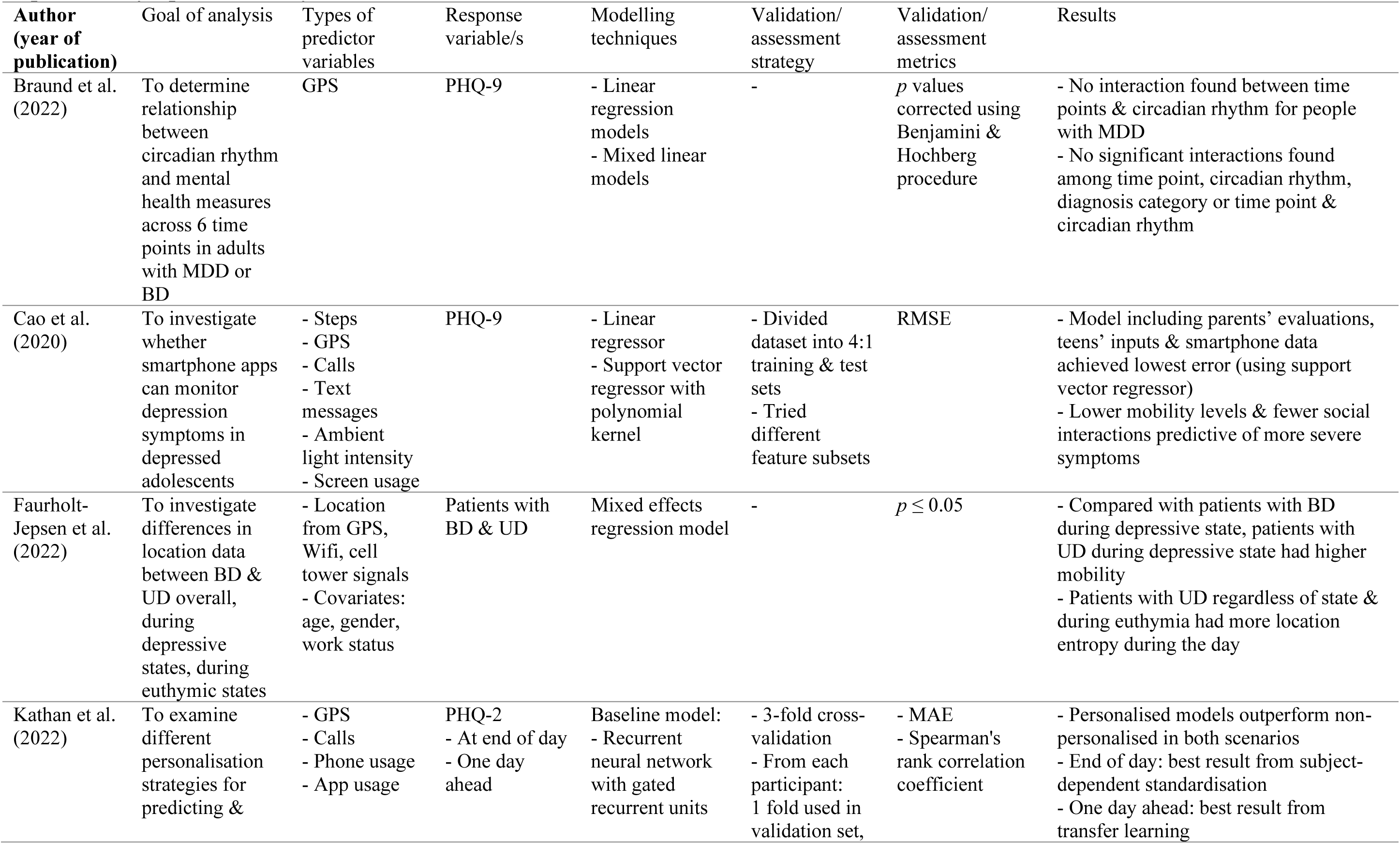
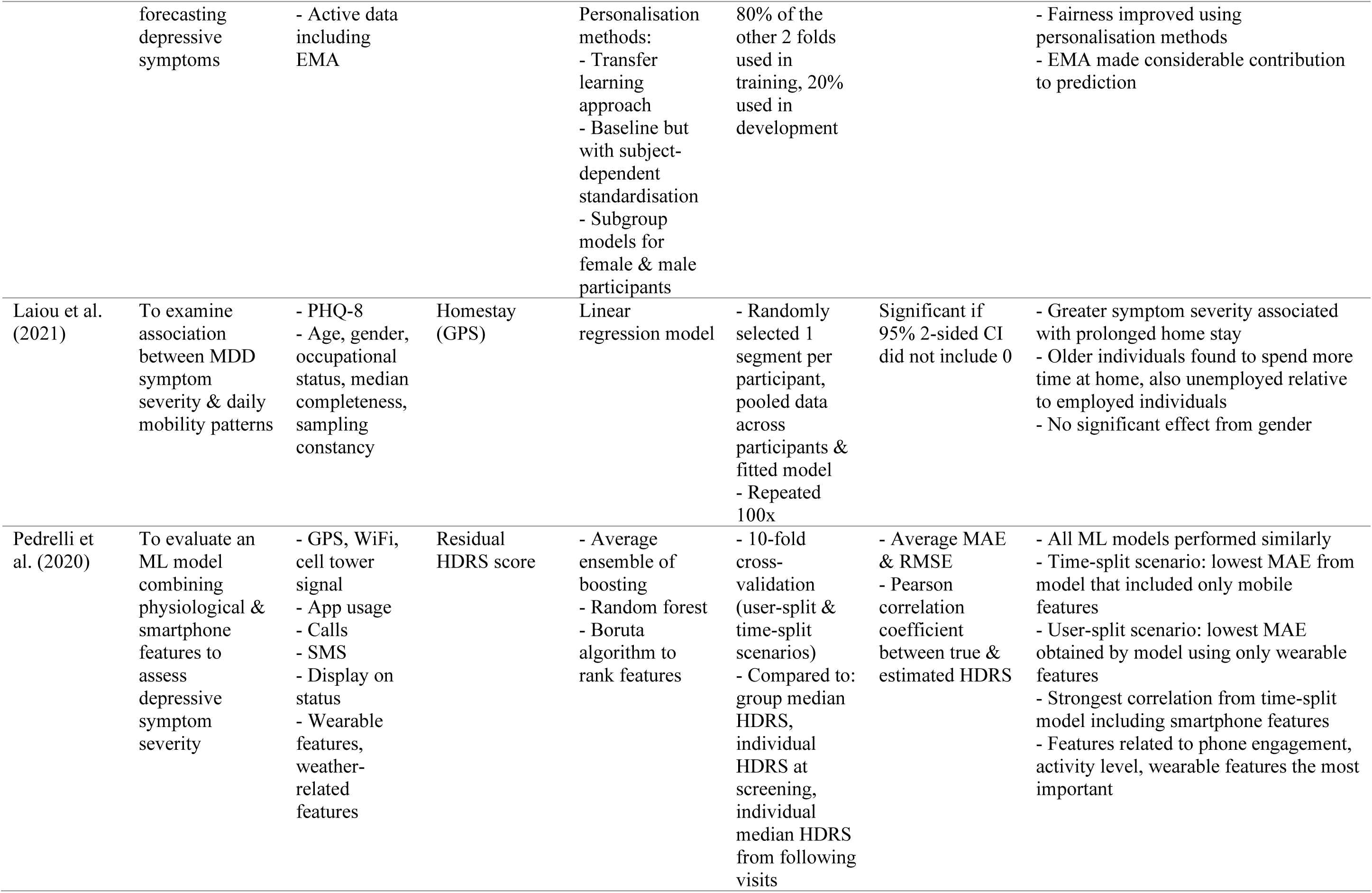
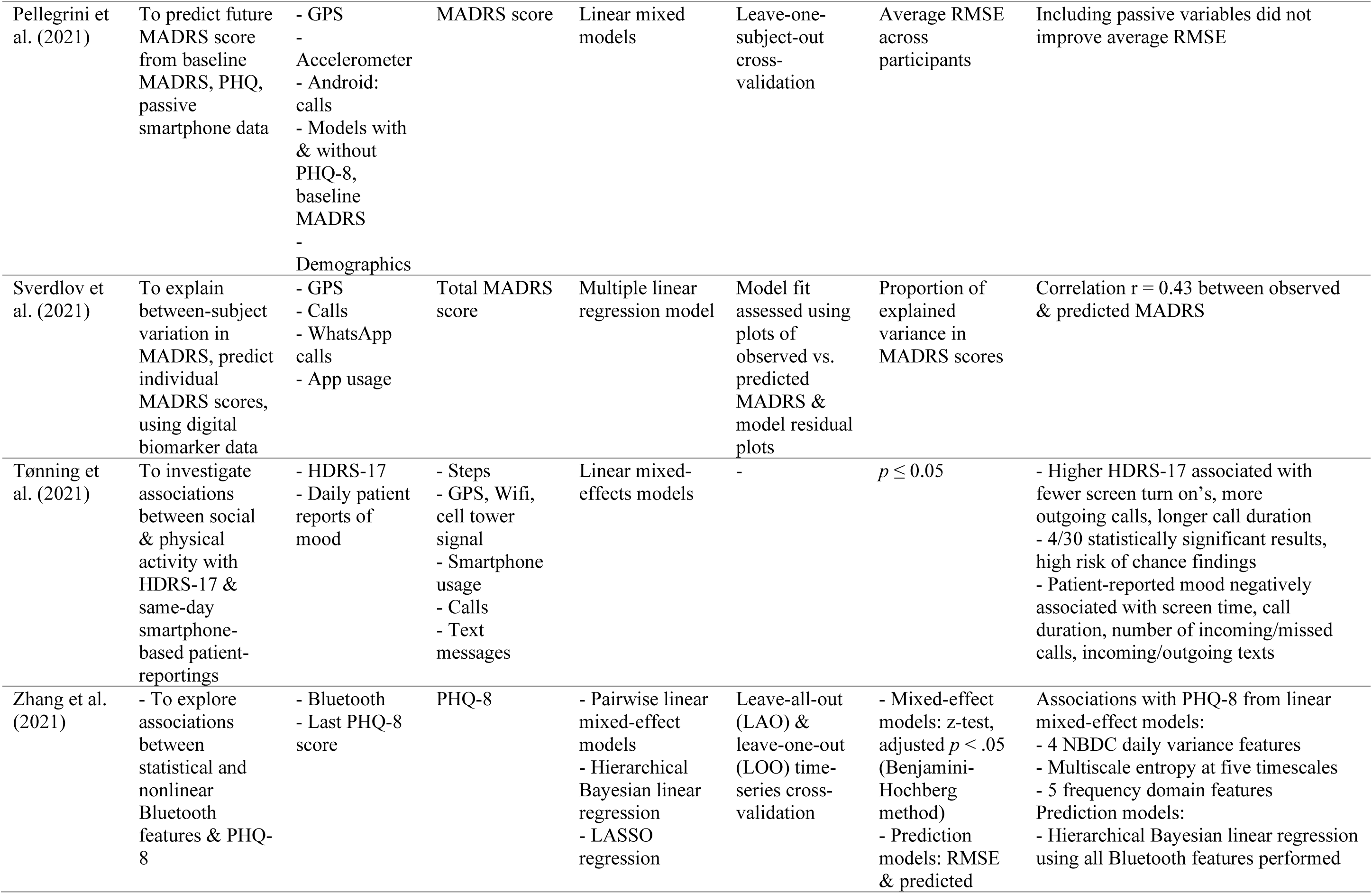
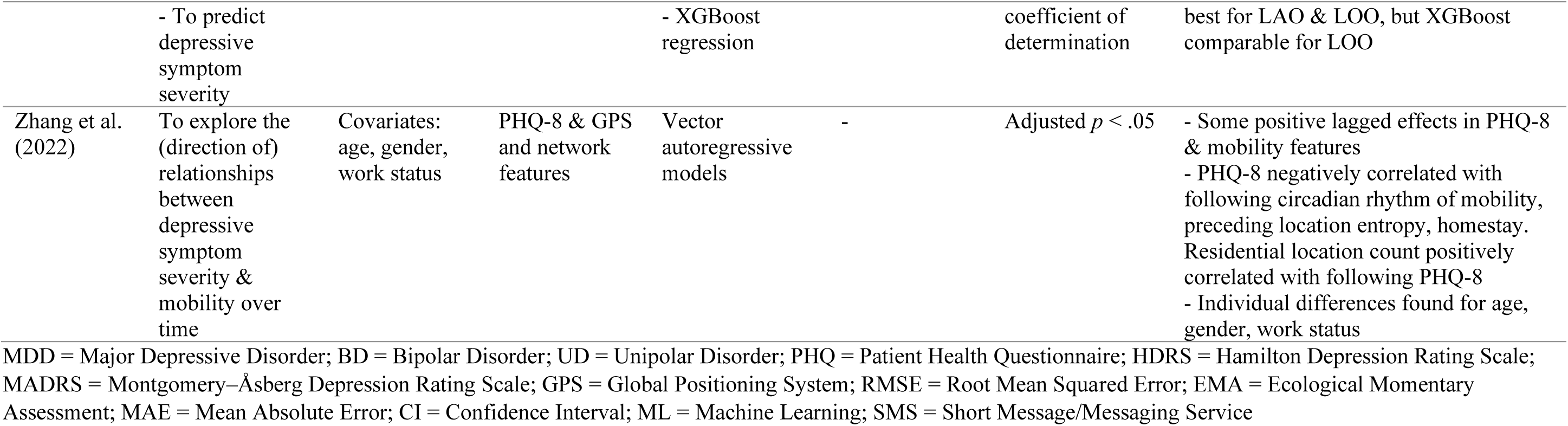
Methodological Details Of Studies Predicting Depression Symptom Severity Or Predicting Passive Smartphone Features From Depression Symptom Severity.

| Author<br>(year of<br>publication) | Goal of analysis | Types of<br>predictor<br>variables | Response<br>variable/s | Modelling<br>techniques | Validation/<br>assessment<br>strategy | Validation/<br>assessment<br>metrics | Results |
| --- | --- | --- | --- | --- | --- | --- | --- |
| Braund et al.<br>(2022) | To determine relationship between circadian rhythm and mental health measures across 6 time points in adults with MDD or BD | GPS | PHQ-9 | - Linear regression models<br>- Mixed linear models | - | *p* values corrected using Benjamini & Hochberg procedure | - No interaction found between time points & circadian rhythm for people with MDD<br>- No significant interactions found among time point, circadian rhythm, diagnosis category or time point & circadian rhythm |
| Cao et al.<br>(2020) | To investigate whether smartphone apps can monitor depression symptoms in depressed adolescents | - Steps<br>- GPS<br>- Calls<br>- Text messages<br>- Ambient light intensity<br>- Screen usage | PHQ-9 | - Linear regressor<br>- Support vector regressor with polynomial kernel | - Divided dataset into 4:1 training & test sets<br>- Tried different feature subsets | RMSE | - Model including parents' evaluations, teens' inputs & smartphone data achieved lowest error (using support vector regressor)<br>- Lower mobility levels & fewer social interactions predictive of more severe symptoms |
| Faurholt-Jepsen et al.<br>(2022) | To investigate differences in location data between BD & UD overall, during depressive states, during euthymic states | - Location from GPS, Wifi, cell tower signals<br>- Covariates: age, gender, work status | Patients with BD & UD | Mixed effects regression model | - | $p \leq 0.05$ | - Compared with patients with BD during depressive state, patients with UD during depressive state had higher mobility<br>- Patients with UD regardless of state & during euthymia had more location entropy during the day |
| Kathan et al.<br>(2022) | To examine different personalisation strategies for predicting & | - GPS<br>- Calls<br>- Phone usage<br>- App usage | PHQ-2<br>- At end of day<br>- One day ahead | Baseline model:<br>- Recurrent neural network with gated recurrent units | - 3-fold cross-validation<br>- From each participant: 1 fold used in validation set, | - MAE<br>- Spearman's rank correlation coefficient | - Personalised models outperform non-personalised in both scenarios<br>- End of day: best result from subject-dependent standardisation<br>- One day ahead: best result from transfer learning |
|  | forecasting depressive symptoms | - Active data including EMA |  | Personalisation methods:<br>- Transfer learning approach<br>- Baseline but with subject-dependent standardisation<br>- Subgroup models for female & male participants | 80% of the other 2 folds used in training, 20% used in development |  | - Fairness improved using personalisation methods<br>- EMA made considerable contribution to prediction |
| Laiou et al. (2021) | To examine association between MDD symptom severity & daily mobility patterns | - PHQ-8<br>- Age, gender, occupational status, median completeness, sampling constancy | Homestay (GPS) | Linear regression model | - Randomly selected 1 segment per participant, pooled data across participants & fitted model<br>- Repeated 100x | Significant if 95% 2-sided CI did not include 0 | - Greater symptom severity associated with prolonged home stay<br>- Older individuals found to spend more time at home, also unemployed relative to employed individuals<br>- No significant effect from gender |
| Pedrelli et al. (2020) | To evaluate an ML model combining physiological & smartphone features to assess depressive symptom severity | - GPS, WiFi, cell tower signal<br>- App usage<br>- Calls<br>- SMS<br>- Display on status<br>- Wearable features, weather-related features | Residual HDRS score | - Average ensemble of boosting<br>- Random forest<br>- Boruta algorithm to rank features | - 10-fold cross-validation (user-split & time-split scenarios)<br>- Compared to: group median HDRS, individual HDRS at screening, individual median HDRS from following visits | - Average MAE & RMSE<br>- Pearson correlation coefficient between true & estimated HDRS | - All ML models performed similarly<br>- Time-split scenario: lowest MAE from model that included only mobile features<br>- User-split scenario: lowest MAE obtained by model using only wearable features<br>- Strongest correlation from time-split model including smartphone features<br>- Features related to phone engagement, activity level, wearable features the most important |
| Pellegrini et al. (2021) | To predict future MADRS score from baseline MADRS, PHQ, passive smartphone data | - GPS<br>- Accelerometer<br>- Android: calls<br>- Models with & without PHQ-8, baseline MADRS<br>- Demographics | MADRS score | Linear mixed models | Leave-one-subject-out cross-validation | Average RMSE across participants | Including passive variables did not improve average RMSE |
| Sverdlov et al. (2021) | To explain between-subject variation in MADRS, predict individual MADRS scores, using digital biomarker data | - GPS<br>- Calls<br>- WhatsApp calls<br>- App usage | Total MADRS score | Multiple linear regression model | Model fit assessed using plots of observed vs. predicted MADRS & model residual plots | Proportion of explained variance in MADRS scores | Correlation $r = 0.43$ between observed & predicted MADRS |
| Tønning et al. (2021) | To investigate associations between social & physical activity with HDRS-17 & same-day smartphone-based patient-reportings | - HDRS-17<br>- Daily patient reports of mood | - Steps<br>- GPS, Wifi, cell tower signal<br>- Smartphone usage<br>- Calls<br>- Text messages | Linear mixed-effects models | - | $p \leq 0.05$ | - Higher HDRS-17 associated with fewer screen turn on's, more outgoing calls, longer call duration<br>- 4/30 statistically significant results, high risk of chance findings<br>- Patient-reported mood negatively associated with screen time, call duration, number of incoming/missed calls, incoming/outgoing texts |
| Zhang et al. (2021) | - To explore associations between statistical and nonlinear Bluetooth features & PHQ-8 | - Bluetooth<br>- Last PHQ-8 score | PHQ-8 | - Pairwise linear mixed-effect models<br>- Hierarchical Bayesian linear regression<br>- LASSO regression | Leave-all-out (LAO) & leave-one-out (LOO) time-series cross-validation | - Mixed-effect models: z-test, adjusted $p < .05$ (Benjamini-Hochberg method)<br>- Prediction models: RMSE & predicted | Associations with PHQ-8 from linear mixed-effect models:<br>- 4 NBDC daily variance features<br>- Multiscale entropy at five timescales<br>- 5 frequency domain features<br>Prediction models:<br>- Hierarchical Bayesian linear regression using all Bluetooth features performed |

|  | - To predict depressive symptom severity |  |  | - XGBoost regression |  | coefficient of determination | best for LAO & LOO, but XGBoost comparable for LOO |
| --- | --- | --- | --- | --- | --- | --- | --- |
| Zhang et al. (2022) | To explore the (direction of) relationships between depressive symptom severity & mobility over time | Covariates: age, gender, work status | PHQ-8 & GPS and network features | Vector autoregressive models | - | Adjusted $p < .05$ | - Some positive lagged effects in PHQ-8 & mobility features<br>- PHQ-8 negatively correlated with following circadian rhythm of mobility, preceding location entropy, homestay. Residential location count positively correlated with following PHQ-8<br>- Individual differences found for age, gender, work status |

MDD = Major Depressive Disorder; BD = Bipolar Disorder; UD = Unipolar Disorder; PHQ = Patient Health Questionnaire; HDRS = Hamilton Depression Rating Scale; MADRS = Montgomery–Åsberg Depression Rating Scale; GPS = Global Positioning System; RMSE = Root Mean Squared Error; EMA = Ecological Momentary Assessment; MAE = Mean Absolute Error; CI = Confidence Interval; ML = Machine Learning; SMS = Short Message/Messaging Service

Applying (penalised) linear regression models to passive smartphone data may help ensure that the models are less likely to overfit to the data, however, the relationship between depression symptom severity and smartphone features may be non-linear. Because of this, several papers chose to investigate predictions of symptom severity by non-linear regression models, often comparing these results to linear regression models. In a study investigating depression in an adolescent population, Cao, et al., (2020) used linear regression and support vector regression to predict PHQ-9 scores from smartphone data and personal and parental ratings. The most accurate model was a support vector regression model combining all three of these data types (RMSE = 2.65). Interestingly, the most accurate model using only smartphone data was a linear model (RMSE = 2.77). Zhang, et al. (2021) used pairwise linear mixed-effect models to explore the relationship between Bluetooth smartphone features and PHQ-8 scores in a cross-country study in populations with a recent history of depression. Features reflected second-order statistics (e.g., the average value of the daily maximum number of nearby Bluetooth device count (NBDC)), multiscale entropy and the frequency domain. Ten of the second-order statistical features, four features related to daily variance of NBDC, multiscale entropy at five timescales and five frequency domain features were associated with depression symptom scores. In general, it was found that for increases in depression symptom severity score, the variance and periodicity of the smartphone features sequence decreased, and it became more irregular. The models containing Bluetooth features provided better fits to the data than a model containing no Bluetooth features. Zhang, et al. (2021) also investigated hierarchical Bayesian linear regression, LASSO regression and XGBoost regression models to predict symptom severity. Their hierarchical Bayesian linear regression model achieved the best performance in terms of their selected metrics for the two different cross-validation scenarios used (RMSE = 3.89, 4.426).

Zhang, et al. (2022) investigated relationships between smartphone features and depression score using vector autoregressive models. They considered (cross-)lagged effects between each time point and the subsequent time point occurring two weeks later. Residential location count was positively correlated with later depression scores (φ = 0.05), despite a negative correlation being found between this feature and depression score at the within-subjects level. Moreover, depressive symptom scores were shown to be negatively correlated with later circadian rhythm (φ = −0.07), and preceding location entropy (φ = −0.04) and homestay (φ = 0.09). Importantly, Zhang, et al. (2022) also demonstrated individual differences in cross-lagged effects related to age and circadian rhythm.

Pedrelli, et al. (2020) aimed to predict HDRS scores using average ensemble of boosting and random forest (AdaBoost) models. These models included smartphone features related to location and movement (using GPS, Wi-Fi and Cell tower signal), app/smartphone usage, and calls/SMS, as well as wearable and weather-related features. Kernel Principal Component Analysis was used to reduce the dimensionality of this feature set from 877 to 25 features, and was also carried out separately for models with only smartphone or wearable features. This study achieved similar performance across models, with the lowest error in a time-split cross-validation scenario from the model including only mobile features (RMSE = 4.88), and the lowest error in a user-split cross-validation scenario from the model including only wearable features (RMSE = 5.35). The machine learning models outperformed predictions made using group median baseline and individual screen baseline models, but not predictions using individual median HDRS scores.

Kathan, et al. (2022) compared personalisation strategies to non-personalised models for prediction of PHQ-2 scores at the end of the day and one day ahead. For end of day prediction, the model using subject-dependent standardisation achieved the best performance (Mean Absolute Error (MAE) = 0.801), and for day ahead prediction the personalisation model using transfer learning achieved the best performance (MAE = 1.349).

Braund, et al. (2022) investigated participants with both bipolar disorder and MDD, using linear regression models to test the association between circadian rhythm and PHQ-9 scores, and mixed-effects linear models to investigate potential moderating effects of circadian rhythm on symptom prediction across six timepoints covering a ten week period. Circadian rhythm was not found to be associated with depression severity and similarly, no interactions were found between time point, circadian rhythm or diagnosis, or time point and circadian rhythm for depression severity. However, there was quite low variability in depression symptom severity indicated graphically in the study, and so strong interactions may be difficult to detect.

Faurholt-Jepsen, et al. (2022) used two-level mixed effects regression models to investigate differences in mobility patterns (quantified using GPS, Wi-Fi and cell tower signals) between participants with bipolar disorder and unipolar depression. During depressive states, participants with unipolar depression were found to cover a significantly larger area per day, and had a larger total distance and duration of moves per day compared to participants with bipolar disorder. Overall and during euthymic states, participants with unipolar depression were found to have greater location entropy during the daytime than participants with bipolar disorder.

### Predicting passive smartphone features

Whilst many studies investigated whether depression symptom severity could be predicted by features derived from passively collected smartphone data, two studies were identified that sought to make the inverse prediction (i.e., predicting different smartphone features from measures of depressive symptoms). Choice of smartphone-based response variables were informed by previous research and clinical knowledge. These studies are displayed in Table 4, alongside the studies in the preceding section. Laiou, et al. (2022) used a linear regression model to predict homestay based on PHQ-8 scores, also including demographic variables in their model (see Table 4). They found that high depression symptom severity was associated with longer home stay during the overall study period and for weekdays only, but not for weekends.

Tønning, Faurholt-Jepsen, Frost, Bardram, and Kessing (2021) investigated the prediction of several smartphone features, including daily averages of physical activity (number of steps, total distance moved), smartphone usage (total screen-on time, number of times screen was turned on) and social activity (number of incoming, outgoing, missed calls, duration of calls, number of incoming & outgoing text messages). Using linear mixed-effects models to account for repeated measurements within each participant, it was found that more severe HDRS scores were significantly associated with fewer screen turn ons, larger number of outgoing calls, and longer phone call durations. However, it was noted that this was a small number of significant results, especially given the high risk of chance findings, as multiple testing was not accounted for. Unlike Zhang, et al. (2022), Tønning, Faurholt-Jepsen, Frost, Bardram, and Kessing (2021) did not find a significant relationship between distance moved and symptom severity. Tønning, Faurholt-Jepsen, Frost, Bardram, and Kessing (2021) also investigated the relationship between the smartphone features and a mood score that patients provided via their smartphones. Lower smartphone-reported mood was associated with increased social activity and phone usage. In this context, incoming communication was suggested to be increased due to concern from external sources.

### Predicting diagnostic class

Studies with classification-related goals are displayed in Table 5. Sverdlov, et al. (2021) investigated two regression methods to classify participants as depressed or healthy. The first was a logistic regression method that utilised input variables (e.g., number of unique places visited, average distance from home, total number of WhatsApp calls, total usage count of apps) selected in a stepwise manner, and for the second method they applied a clinically-determined threshold to MADRS scores predicted by a multiple linear regression model (again using selected input variables) to split the participants into the two classes. The latter model achieved higher accuracy (0.75), sensitivity and area under the receiver operating characteristic curve (AUC) than the logistic regression model, and comparable specificity. Kim, et al., 2023 also classified participants with MDD versus healthy controls, using deep neural networks and SVM. The deep neural network achieved higher accuracy in cross-validation (77%). Additionally. Kim, et al., 2023 classified MDD patients who responded to antidepressant treatment versus those who did not respond, achieving 85% cross-validation accuracy using an SVM.

**Table 5.**
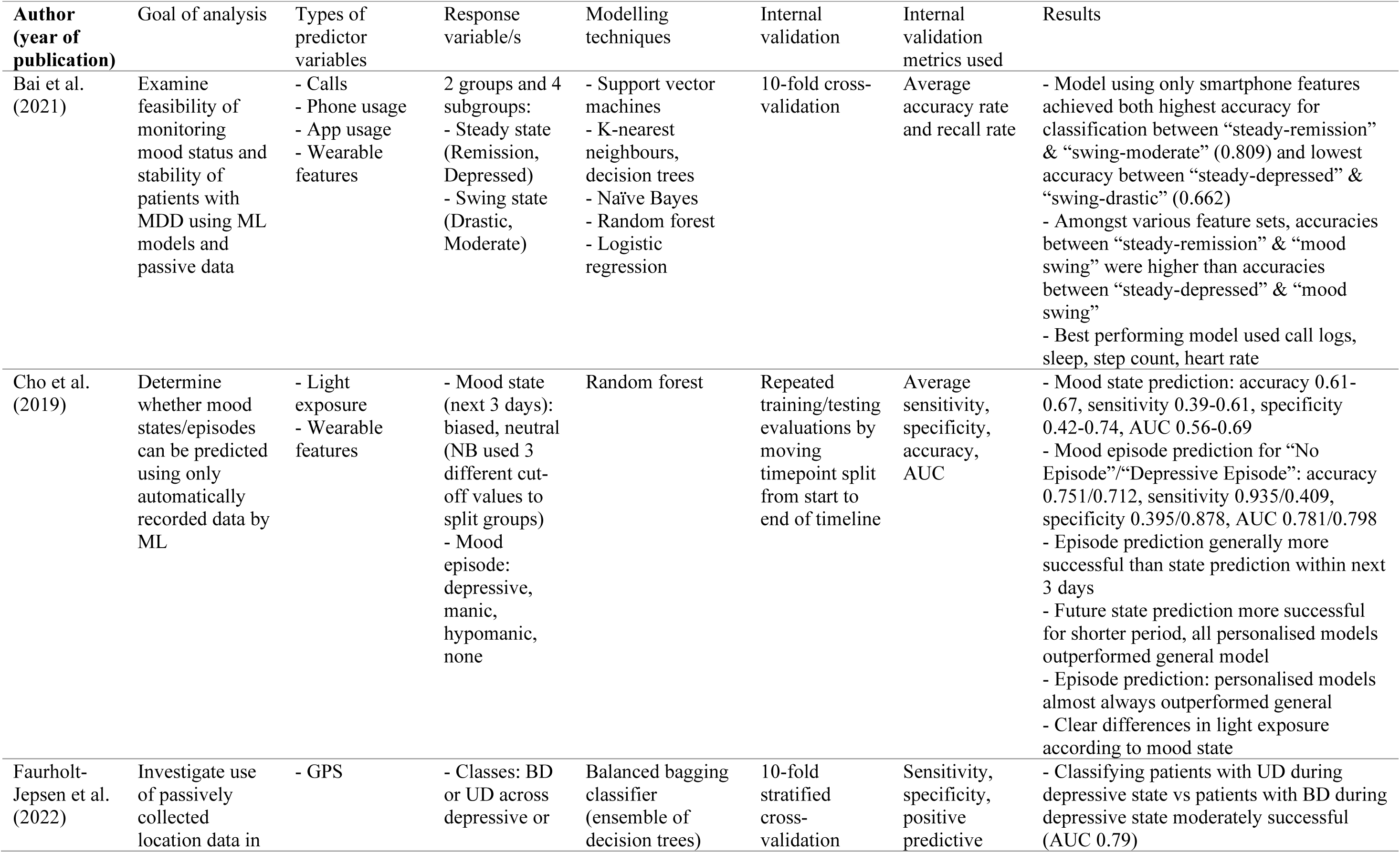
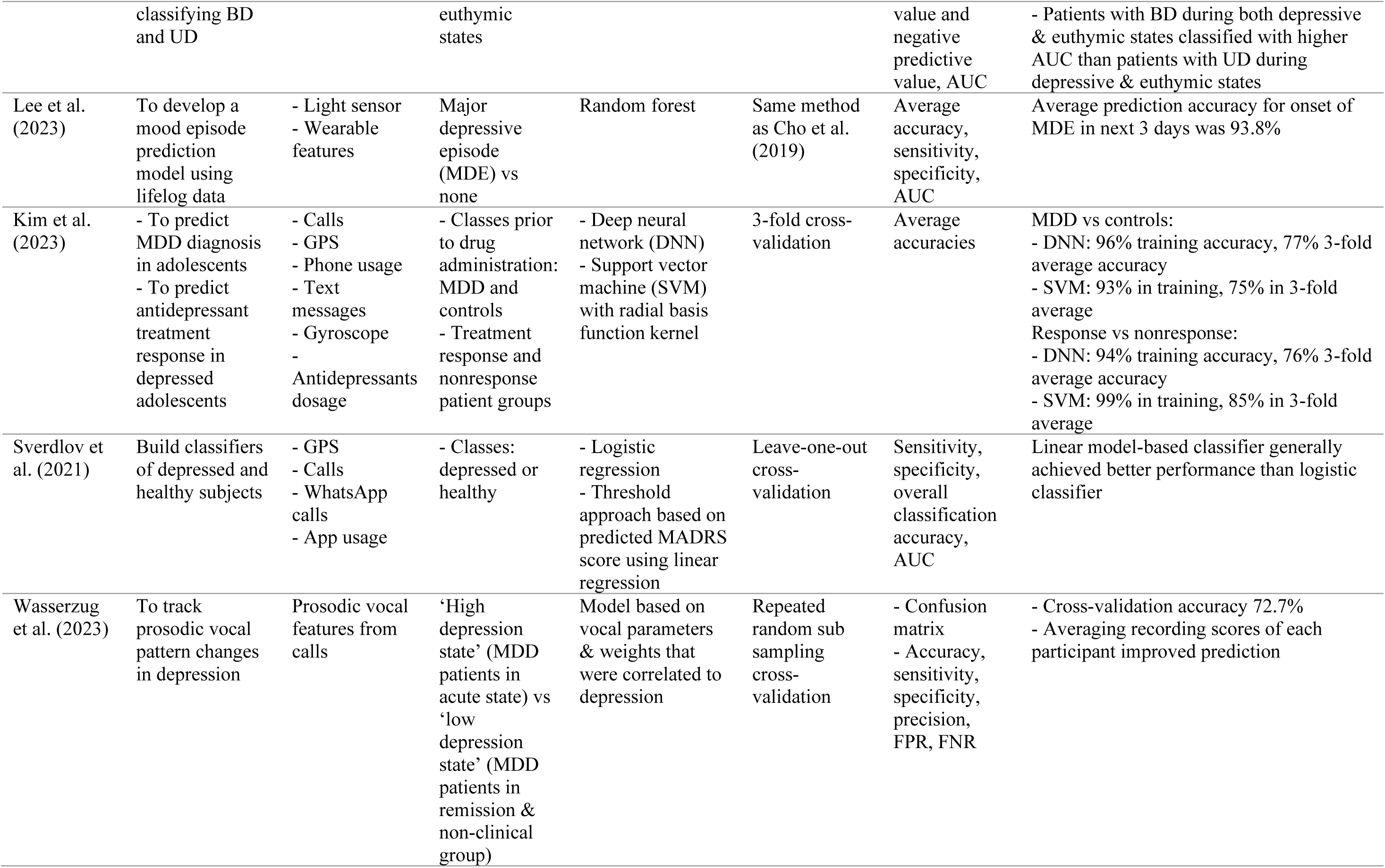
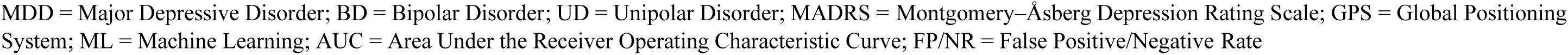
Methodological Details Of Studies Predicting Diagnostic Class Or Mood State/Episode Label.

| Author<br>(year of<br>publication) | Goal of analysis | Types of<br>predictor<br>variables | Response<br>variable/s | Modelling<br>techniques | Internal<br>validation | Internal<br>validation<br>metrics used | Results |
| --- | --- | --- | --- | --- | --- | --- | --- |
| Bai et al.<br>(2021) | Examine feasibility of monitoring mood status and stability of patients with MDD using ML models and passive data | - Calls<br>- Phone usage<br>- App usage<br>- Wearable features | 2 groups and 4 subgroups:<br>- Steady state (Remission, Depressed)<br>- Swing state (Drastic, Moderate) | - Support vector machines<br>- K-nearest neighbours, decision trees<br>- Naïve Bayes<br>- Random forest<br>- Logistic regression | 10-fold cross-validation | Average accuracy rate and recall rate | - Model using only smartphone features achieved both highest accuracy for classification between “steady-remission” & “swing-moderate” (0.809) and lowest accuracy between “steady-depressed” & “swing-drastic” (0.662)<br>- Amongst various feature sets, accuracies between “steady-remission” & “mood swing” were higher than accuracies between “steady-depressed” & “mood swing”<br>- Best performing model used call logs, sleep, step count, heart rate |
| Cho et al.<br>(2019) | Determine whether mood states/episodes can be predicted using only automatically recorded data by ML | - Light exposure<br>- Wearable features | - Mood state (next 3 days): biased, neutral (NB used 3 different cut-off values to split groups)<br>- Mood episode: depressive, manic, hypomanic, none | Random forest | Repeated training/testing evaluations by moving timepoint split from start to end of timeline | Average sensitivity, specificity, accuracy, AUC | - Mood state prediction: accuracy 0.61-0.67, sensitivity 0.39-0.61, specificity 0.42-0.74, AUC 0.56-0.69<br>- Mood episode prediction for “No Episode”/“Depressive Episode”: accuracy 0.751/0.712, sensitivity 0.935/0.409, specificity 0.395/0.878, AUC 0.781/0.798<br>- Episode prediction generally more successful than state prediction within next 3 days<br>- Future state prediction more successful for shorter period, all personalised models outperformed general model<br>- Episode prediction: personalised models almost always outperformed general<br>- Clear differences in light exposure according to mood state |
| Faurholt-Jepsen et al.<br>(2022) | Investigate use of passively collected location data in | - GPS | - Classes: BD or UD across depressive or | Balanced bagging classifier (ensemble of decision trees) | 10-fold stratified cross-validation | Sensitivity, specificity, positive predictive | - Classifying patients with UD during depressive state vs patients with BD during depressive state moderately successful (AUC 0.79) |

|  | classifying BD and UD |  | euthymic states |  |  | value and negative predictive value, AUC | - Patients with BD during both depressive & euthymic states classified with higher AUC than patients with UD during depressive & euthymic states |
| --- | --- | --- | --- | --- | --- | --- | --- |
| Lee et al. (2023) | To develop a mood episode prediction model using lifelog data | - Light sensor<br>- Wearable features | Major depressive episode (MDE) vs none | Random forest | Same method as Cho et al. (2019) | Average accuracy, sensitivity, specificity, AUC | Average prediction accuracy for onset of MDE in next 3 days was 93.8% |
| Kim et al. (2023) | - To predict MDD diagnosis in adolescents<br>- To predict antidepressant treatment response in depressed adolescents | - Calls<br>- GPS<br>- Phone usage<br>- Text messages<br>- Gyroscope<br>- Antidepressants dosage | - Classes prior to drug administration: MDD and controls<br>- Treatment response and nonresponse patient groups | - Deep neural network (DNN)<br>- Support vector machine (SVM) with radial basis function kernel | 3-fold cross-validation | Average accuracies | MDD vs controls:<br>- DNN: 96% training accuracy, 77% 3-fold average accuracy<br>- SVM: 93% in training, 75% in 3-fold average<br>Response vs nonresponse:<br>- DNN: 94% training accuracy, 76% 3-fold average accuracy<br>- SVM: 99% in training, 85% in 3-fold average |
| Sverdlov et al. (2021) | Build classifiers of depressed and healthy subjects | - GPS<br>- Calls<br>- WhatsApp calls<br>- App usage | - Classes: depressed or healthy | - Logistic regression<br>- Threshold approach based on predicted MADRS score using linear regression | Leave-one-out cross-validation | Sensitivity, specificity, overall classification accuracy, AUC | Linear model-based classifier generally achieved better performance than logistic classifier |
| Wasserzug et al. (2023) | To track prosodic vocal pattern changes in depression | Prosodic vocal features from calls | 'High depression state' (MDD patients in acute state) vs 'low depression state' (MDD patients in remission & non-clinical group) | Model based on vocal parameters & weights that were correlated to depression | Repeated random sub sampling cross-validation | - Confusion matrix<br>- Accuracy, sensitivity, specificity, precision, FPR, FNR | - Cross-validation accuracy 72.7%<br>- Averaging recording scores of each participant improved prediction |

MDD = Major Depressive Disorder; BD = Bipolar Disorder; UD = Unipolar Disorder; MADRS = Montgomery–Åsberg Depression Rating Scale; GPS = Global Positioning System; ML = Machine Learning; AUC = Area Under the Receiver Operating Characteristic Curve; FP/NR = False Positive/Negative Rate

**Table 6.**
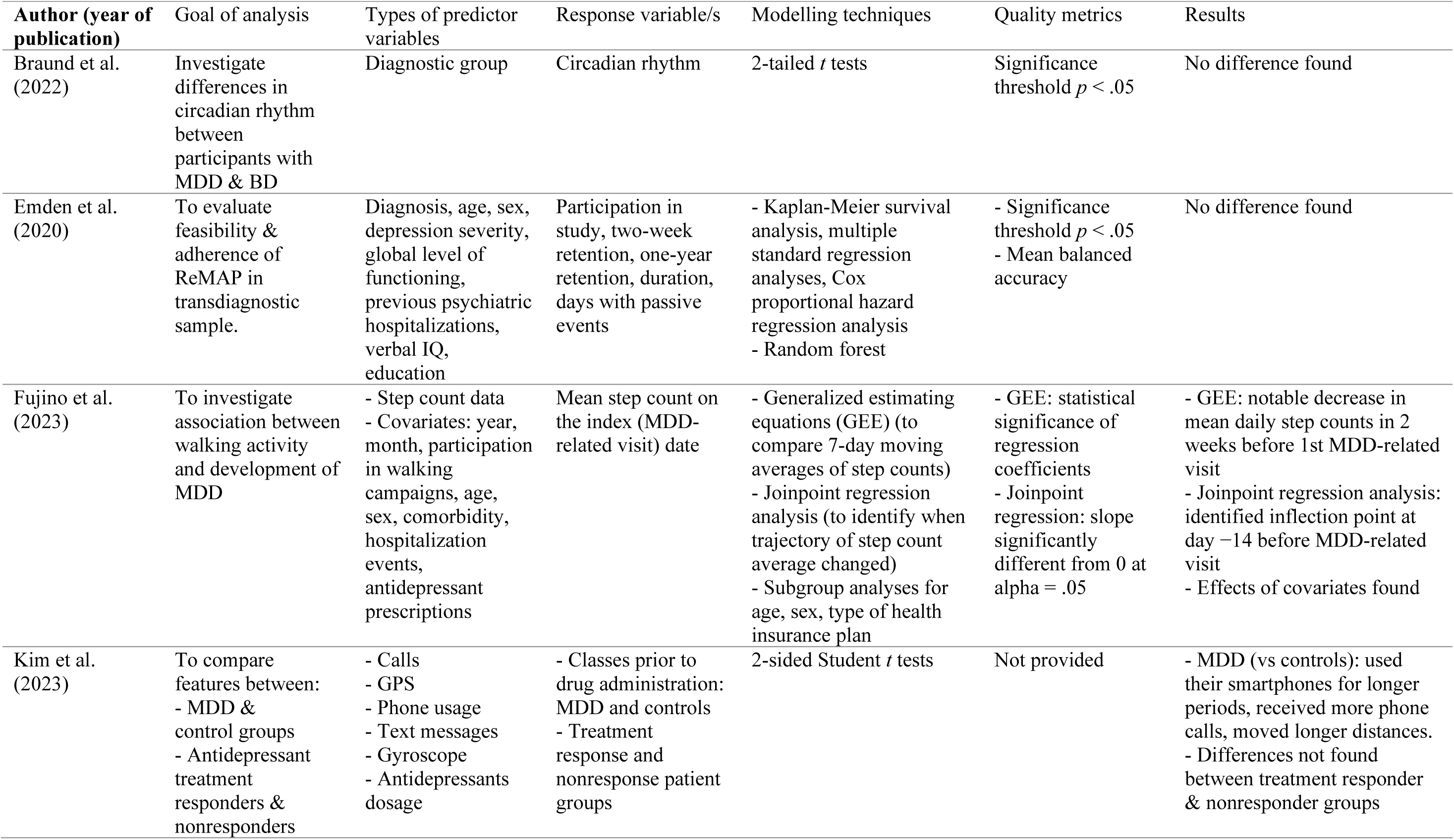
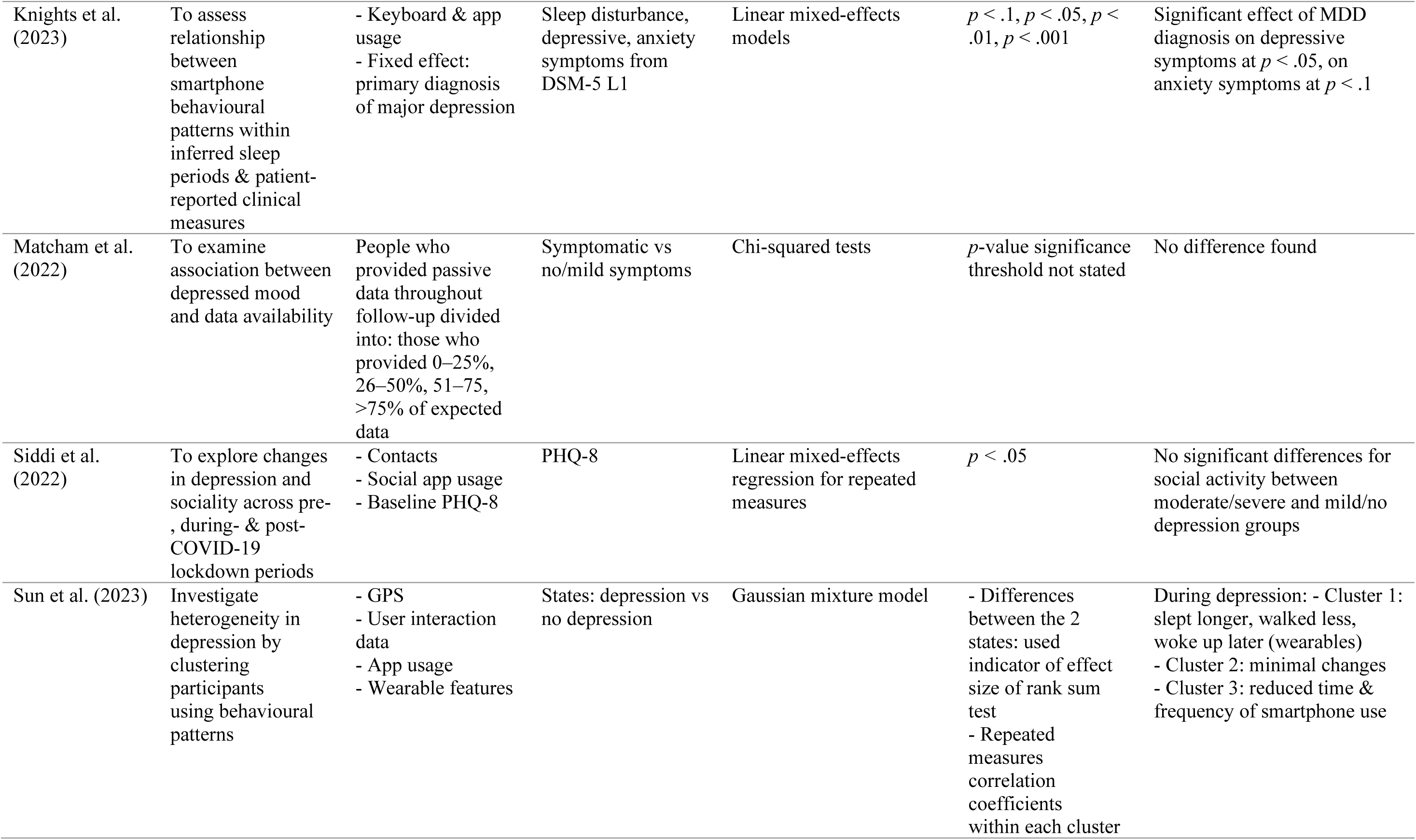
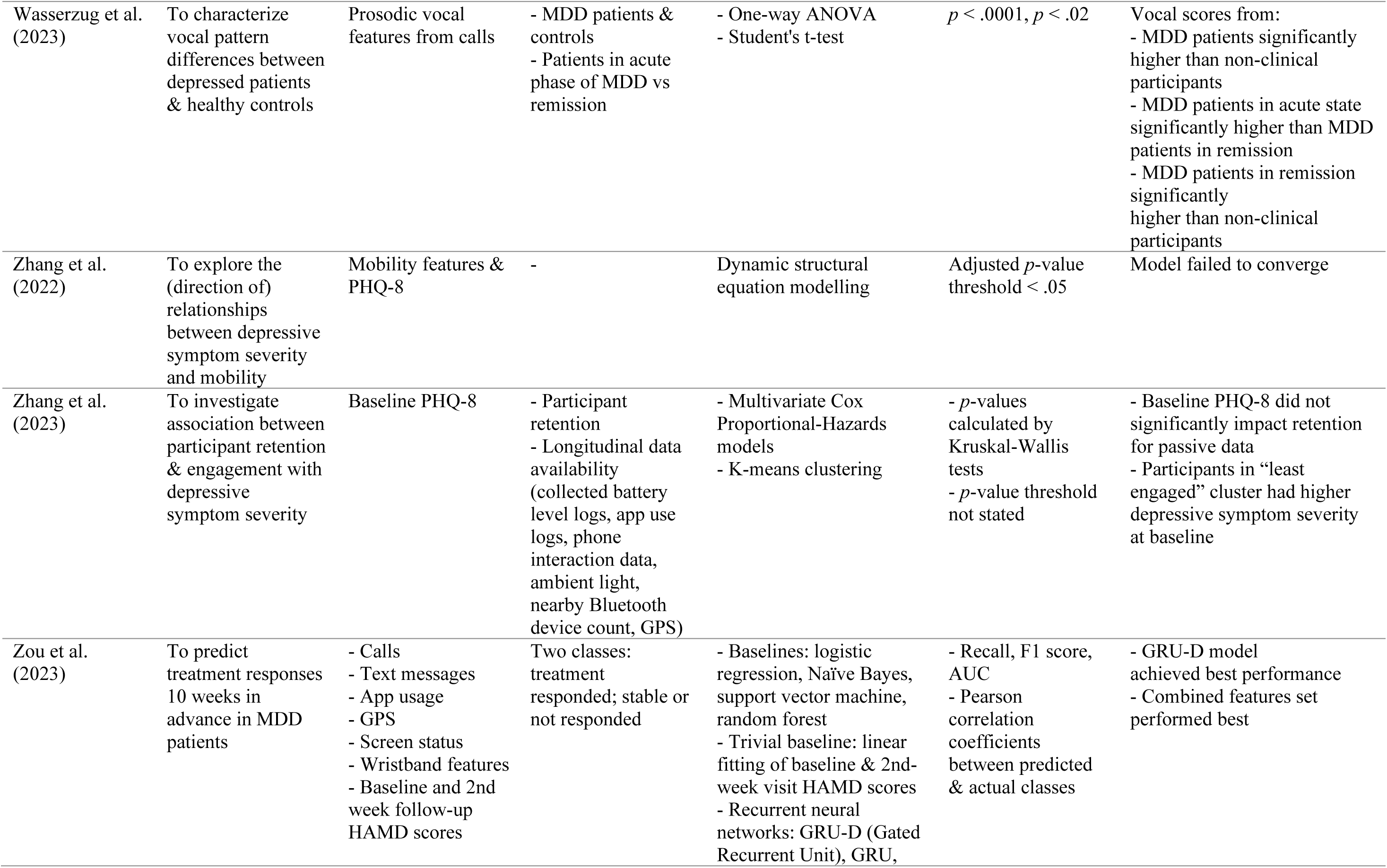
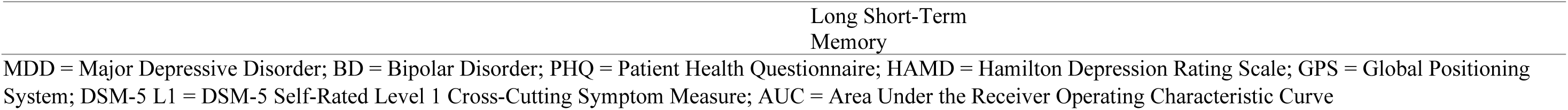
Methodological Details Of Studies With Other Analysis Goals.

Faurholt-Jepsen, et al. (2022) classified participants with bipolar disorder and unipolar depression into overall diagnostic classes using an ensemble of decision trees. The model achieved a sensitivity of 0.70, specificity of 0.65, and AUC of 0.75 during cross-validation. In line with this, Faurholt-Jepsen, et al. (2022) aimed to classify solely the depressive periods of participants with bipolar disorder or unipolar depression, to investigate whether the depressive state of participants in the two diagnostic groups can be differentiated, achieving again a sensitivity of 0.70, a higher specificity of 0.77, and AUC of 0.79. Overall, it can be seen that these models could differentiate between classes with moderate success, bearing in mind that a random binary classifier would achieve about 50% accuracy.

### Predicting mood state/episode label

Rather than focusing on diagnostic labels, many studies chose to focus on mood states (these studies are also included in Table 5). This approach may be a useful step towards predicting clinically relevant changes in state for those who already have a diagnosis, or to predict relapse for those in remission. Bai, et al. (2021) aimed to classify participants with MDD into two groups, steady state and mood swing state, as well as four subgroups (steady state: in remission, currently depressed; mood swing: drastic (i.e., difference between maximum and minimum PHQ-9 scores is greater than or equal to ten), moderate (i.e., difference in scores is greater than or equal to five)), using a variety of machine learning methods. Statistical smartphone features were calculated for the different types of phone calls and times of call, call duration, number of people involved in the calls and the entropy of callers. Some features from wearable devices (e.g., step count, heart rate) were also used. The success rate of classification was found to vary depending on which classes and features were included in each binary classifier. The set of features that generally gave models with the highest accuracies contained features related to call logs, sleep data, step count data, and heart rate data. The models using features from only smartphone data generally achieved the lowest accuracies. Although Bai et al. (2021) chose to focus on binary classifiers, a multi-class classifier may have more clinical utility as in the real world the class label would not yet be known, therefore it would be uncertain which binary classifier would be appropriate.

Wasserzug, et al. (2023) classified MDD patients (in acute depressive states and in remission) and healthy controls into ‘high depression’ and ‘low depression’ states using a model developed from vocal features that were correlated with depression, achieving a cross-validation accuracy of 72.7%.

Random forest models were used by Cho, et al. (2019) to classify mood state as biased or neutral, and mood episode as depressive, manic, hypomanic, or no episode. Behavioural patterns from participants with bipolar disorder were also investigated. Cho, et al. (2019) used statistical features related to light exposure during bedtime and daytime, and many Fitbit features. For the mood state prediction of patients with MDD, and various mood score cut-off values, the accuracy ranged from 0.61-0.67. For the mood episode prediction of patients with MDD, the accuracy was 0.751 and 0.712 for “No Episode” and “Depressive Episode” respectively. Cho, et al. (2019) also investigated individual models of mood state and episode classification, finding that the personalised mood state model outperformed the general model in all cases. For mood episode prediction, the personalised model achieved better performance in almost all cases.

#### Mood state prediction at future time points

The mood state models reported in Cho, et al. (2019) involved making predictions related to future time points. That is, Cho, et al. (2019) used a classifier to predict mood states three days following the window covered by the data collection of passive smartphone features, using a mixture of smartphone-derived light exposure features and wearable features. The number of days used to test the model (i.e., the three days) and the number of days used to train the model (18 days) were selected during parameter tuning, with longer periods (up to 300 days for training days and 30 days for testing) also being investigated. However, the shorter period of three days was found to be a more reasonable window for mood state prediction than longer periods during their parameter selection process, suggesting that later mood state prediction was difficult. In a follow-up study, Lee, et al., 2023 classified the presence or absence of major depressive episodes in the next three days in MDD patients, achieving an average prediction accuracy of 93.8% in their validation method. The prediction method used in Cho, et al. (2019) and Lee, et al., 2023 differed from other studies that aimed to make predictions for mood at the end of smartphone data collection (e.g., Pellegrini, et al., 2022), and gives an example of how digital phenotyping research can shift towards predicting upcoming depression states.

### Other analysis goals

A few studies investigated digital phenotyping in participants with MDD but could not be categorised into one of the above groups, or contained analyses that could not be categorised into these groups. For completeness, these studies are summarised in Table 6 and selected analyses (that were particularly relevant to clinical applications) reported here. Our decision to present analyses in this section was also informed by whether the overall study had already had key analyses addressed in earlier sections of the Results. Fujino, Tokuda, & Fujimoto (2023) investigated changes in smartphone step count surrounding MDD-related medical visits in a group-based analysis, finding that mean daily step count tended to decrease in the two weeks before the visits. Zou, et al., 2023 predicted treatment response in MDD patients 10 weeks in advance, achieving an AUROC (Area Under the Receiver Operating Characteristic Curve) of 0.65. Braund, et al. (2022) investigated differences in circadian rhythm between participants with MDD and participants with bipolar disorder, not identifying a difference between groups.

Emden, Goltermann, Dannlowski, Hahn, and Opel (2021) investigated differences in study participation or retention between various diagnostic groups (affective, anxiety, and psychotic disorder groups and healthy controls), which yielded no significant differences between groups. Focusing on participants with MDD, Matcham, et al. (2022) investigated whether depressed mood was associated with data availability, not identifying a difference in data availability between those with no or mild depressive symptoms and more severe symptoms. In a later study from this consortium, Zhang, et al., 2023 also did not find a significant impact of baseline depression score on participant retention for passive data, however did now find differences in data availability.

### Comparison of Study Methods

#### Feature construction

Due to the vast range of sensors on smartphones, there are many different options of feature sets that are available or chosen for digital phenotyping. Sensors used by the studies identified in this review included, for example, GPS, light, steps, app data, smartphone on-off status, Wi-Fi and Bluetooth. The sensor data can be processed in many ways to create features, for example duration, count and statistical features. The number of studies using the various feature types are displayed in Figure 2.

**Figure 2.**
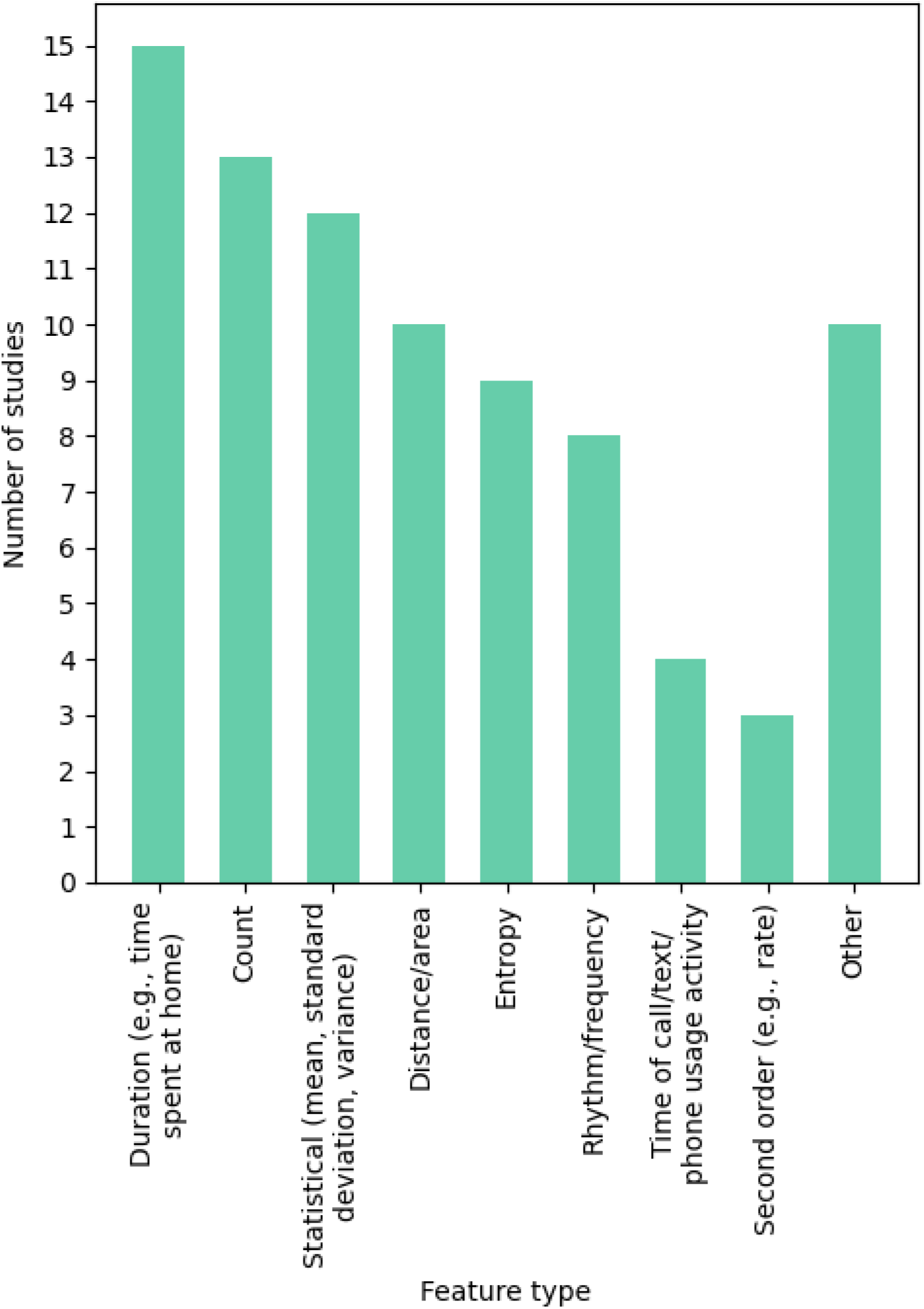
Number of studies (out of the 24 selected studies) using each broad category of feature type (N.B., studies generally used more than one feature type).

Several different processing steps were used in the calculation of features (see Table 2). Clustering was sometimes used to group GPS samples into separate locations, for example by using the Density-based spatial clustering of applications with noise (DBSCAN) algorithm (Faurholt-Jepsen, et al., 2022; Kathan, et al., 2022; Sun, et al., 2023; Zhang, et al., 2022) or K-means clustering (Cao, et al., 2020; Kathan, et al., 2022). Various thresholds were also used, for example to set minimum requirements for clusters. Various requirements were used in the identification of a home location. Several studies chose to use dimension reduction or feature selection methods to reduce the number of features in their feature set/s, including Principal Component Analysis (Pellegrini, et al., 2022; Pedrelli, et al., 2020), L1-Based Feature Selection and Tree-Based Feature Selection (Bai, et al., 2021), feature selection using neural networks (Kim, et al., 2023) and Principal Feature Analysis (Sun, et al., 2023).

### Handling of missing and invalid data

A common issue amongst the studies in this review was the prevalence of missing smartphone data. Various tactics were used to handle this issue (see Table 2). Some studies did not mention any strategy for handling missing data, whilst others did not explicitly indicate the strategy used to deal with this, but acknowledge some kind of criteria. For example, by mentioning the use of “rich sensor data” (Cao, et al., 2020), or stating that analyses were only performed when sufficient data was deemed to be available (Braund, et al., 2022). Other studies explicitly stated thresholds for data inclusion (Bai, et al., 2021; Faurholt-Jepsen, et al., 2022; Fujino, Tokuda, & Fujimoto, 2023; Kathan, et al., 2022; Knights, Shen, Mysliwiec, & DuBois, 2023; Laiou, et al., 2022; Sun, et al., 2023; Zhang, et al., 2021; Zhang, et al., 2022; Zou, et al., 2023); if too high a percentage of data for a sample was missing then the sample was excluded. There was no consistent selection of thresholds for data inclusion across studies.

In other studies, researchers chose to exclude days that had any variable missing (Pellegrini, et al., 2022; Pedrelli, et al., 2020; Cho, et al., 2019). However, Cho, et al. (2019) indicated that not being able to incorporate missing data in models can lead to loss of large volumes of data, as illustrated by the exclusion of approximately 89% of their samples due to the inability of the selected model to account for missing data. Occasionally imputation was carried out on missing data (Kathan, et al., 2022; Lee, et al., 2023; Pedrelli, et al., 2020; Pellegrini, et al., 2022; Zhang, et al. 2021), usually in combination with a minimum criterion of available data.

Smartphone measures may also contain invalid data, for example implausible outlier values. The handling of outliers was not consistently addressed and in many cases not referred to (see Table 2), although it was more common for this to be addressed in studies using location data by excluding unrealistic points (Faurholt-Jepsen, et al., 2022; Laiou, et al., 2022; Sun, et al., 2023; Zhang, et al., 2022). The optimal strategy for handling outliers is highly dependent on the types of measures that are included as well as their distributions, further limiting the comparability across studies.

## Discussion

MDD is a very common and debilitating mental disorder, associated with a high recurrence risk. The recent decade witnessed an upsurge of digital phenotyping studies to better understand dynamics of MDD and to advance precision methods efforts for (relapse) prevention. This systematic review investigated the different features and methods used in smartphone-based digital phenotyping research for MDD and their associated predictive power, identifying a total of 24 studies. These studies overall supported the use of digital phenotyping in MDD, but conjunctively showed that the field is still in relatively early stages, with much room for improvement in predictive performance and the understanding of individual differences in digital phenotypes still to be more rigorously developed. Six of the studies investigated correlations between smartphone-derived features and depressive symptoms, and nine studies predicted depressive symptom scores from passive smartphone data (or in two studies, the inverse prediction). Seven studies also sought to predict classes of data, including diagnosis, mood state or mood episode, with two studies predicting mood state three days in advance (Cho, et al., 2019; Lee, et al., 2023).

Our first research question aimed to establish which passive smartphone features are correlated with clinically relevant variables in MDD. Features generally fell into three broad categories: communication, phone usage and mobility. Despite differences in the specific features, meaning that few similar features were calculated in more than one study, common themes arose: higher depression symptom scores were associated with lower mobility and social interaction measures.

Our second research question aimed to investigate the different methods that have been used for depression prediction tasks using smartphone data, and their performance. For predicting depressive symptom severity, linear (mixed) models were commonly used, with nonlinear methods less common. To predict classes of diagnosis, mood state and mood episode, classic classification algorithms such as random forest and support vector machines were popular choices. Overall, studies tended to achieve moderate predictive performance. For example, with the exception of one classification study achieving over 90% accuracy in the internal validation procedure (Lee, et al., 2023), the highest out-of-sample classification accuracies achieved during cross-validation tended to be in the high 70s to low 80s. Lower accuracies were frequently reported, indicating relatively varied predictive value of current smartphone-based digital phenotyping methods. Moreover, few studies aimed to predict responses across time points. Of note, Fujino, Tokuda, & Fujimoto (2023) carried out a time-resolved analysis that investigated changes in step count surrounding MDD-related medical visits. Finally, a small number of studies investigated individual differences, demonstrating differences between participants of different ages and employment statuses (e.g., in time spent at home (Zhang, et al., 2022; Laiou, et al., 2022)), as well as some gender differences (e.g., in location entropy and residential location count (Zhang, et al., 2022)). This is consistent with the common intuition that individuals have different phone use and behavioural habits, and suggests that factors such as these could be included in prediction models to improve personalised predictions.

In the following sections, we describe key methodological themes identified in this review, and discuss the importance of feature construction in digital phenotyping. We then discuss important study differences that affect the comparison between studies, and identify limitations of this review, before discussing recommendations for the field of digital phenotyping.

### Key methodological themes

The richness and complexity of digital phenotyping data brings about several challenges that need to be overcome through making careful methodological decisions. Due to the high temporal resolution of the data and many available feature options that contribute to high heterogeneity of the data, many studies chose to include time-averaged features to summarise their data. This is a practical way to reduce the large volumes of data to a smaller, more manageable, number of features. However, depending on the chosen level of granularity, this approach can greatly reduce information relating to the temporal dynamics of individual time series. Inclusion of measures reflecting variations in rhythm may have been done to overcome this disadvantage. For some features, the duration of data collection may affect their utility. For example, features such as circadian rhythm may be more reliably calculated when data is collected over a longer period of time, as this may provide indications of the relevance of variations given a person’s “usual” behaviours. As many studies focused on shorter periods (e.g., two week windows), future studies may seek to focus on changes in digital phenotypes over longer periods of time, such as in the range of months/years rather than weeks. Of note, approximately half of the studies involved follow-up/longitudinal clinical data, with the remainder either not collecting follow-up data, or not reflecting this element of study design in the subsequent analysis (see Table 2). Studies also generally validated time-averaged digital phenotyping measures against symptom measures that are also time-averaged, for example by comparing averages from two weeks of digital phenotyping data to a PHQ score reflecting depressive symptoms over two weeks. Whilst this is a necessary step during the early stages of digital phenotyping, it may detract from one of the original motivations behind digital phenotyping; i.e., to develop tools that can be used for real-time patient monitoring.

In this review we use the term “external validation” to refer to the use of datasets that are independent from the main analysis dataset, for example datasets that were collected through a separate study, to evaluate model performance. We do not consider withheld data from the same dataset to be external validation due to a lack of independence. None of the included studies used external validation datasets to assess model performance, with results either reflecting the overall dataset, or an internal validation dataset (i.e., a subset of the overall dataset). Methods such as k-fold cross-validation help to give an indication of performance on withheld data, but do increase the risk of bias. External validation/testing sets may be difficult to obtain due to limited data availability, which could be aided in the future through more cross-collaboration and sharing of datasets.

High prevalence of missing data is a serious concern in smartphone-based digital phenotyping research, as the various different functionalities of smartphones may be unable to function under certain conditions (e.g., low battery, poor Wi-Fi connection, or intended smartphone switch-offs by users). Strategies for handling missing data therefore need to be applied. Studies identified by this review used minimum data inclusion criteria, exclusion of samples with missing values and/or imputation of missing values. The latter two options allow flexibility in the chosen models as they do not need to be able to account for missing data. However, as data may be missing due to the behaviours of the participants (e.g., missingness as a result of an ongoing depressive episode), it is possible that imputing missing data may disguise important behavioural changes. For example, Cohen, et al. (2023) found that their “data quality” feature, calculated based on the data that was successfully collected relative to the amount of data that was expected to be collected, was useful in predicting relapse in schizophrenia. This may also be the case for predicting clinically significant changes in depression. One study included in this review also investigated the impact of missing data on feature quality (Sun, et al., 2023), finding that a range of days’ worth of data (2-12 of 14 days) are needed to reliably calculate different features. Results such as these could inform the selection of missing data thresholds for different feature types.

### Feature construction

As a large number of available sensors and options exist that researchers can select from before data collection begins, appropriate feature construction is key in digital phenotyping. This feature construction seems particularly guided by the technical availability of specific sensors and options, as different smartphone operating systems (e.g., Android, IoS) and applications have various restrictions. For example, Apple smartphones tend to have more restrictions on what data can be collected, and in applications such as WhatsApp, information about calls and messages cannot be accessed. Besides “technically-driven feature construction”, concerns about ethics and/or privacy (e.g., Maher, et al., 2019) can also drive the choice of features. For example, no study analysed the predictive value of content of general phone text messages, and only one study utilised voice recordings (Wasserzug, et al., 2023). Passive voice collection is likely less popular due to ethical issues and privacy concerns, although active voice recordings have shown promise in differentiating between people with depression and healthy controls (Silva, Lopes, Galdino, & Almeida, 2021). Of the excluded studies, some did record voices in defined settings or through using specific exercises or assessments (e.g., Abbas, et al., 2021).

Moreover, selected feature sets tended to be influenced by domain knowledge of depression symptoms (“theory-driven feature construction”). For example, measures of home stay were common, and many studies included measures of behavioural rhythm, such as circadian rhythm (see Figure 2). Two studies (Cao, et al., 2020; Kathan, et al., 2022) included a proxy measure of sleep in their prediction models, although with known limitations as it is not expected that participants use their smartphones immediately before and after sleep. Relatedly, more “data-driven” feature constructions were also seen to be used, in which a broad range of features are calculated to investigate which are most useful (e.g., statistical features calculated for a measure of movement). There are also other features that seem to fall between theory- and data-driven; for example, screen time is a commonly used feature yet despite its common perception, it may not have a notable negative impact on mental health (Aschbrenner, et al., 2019). As we do not yet fully understand the impact of this behaviour on mental health, its use as a digital marker cannot be considered to be completely theory-driven. Studies also did not record reasons a person may have for greater screen time (e.g., family or work commitments), so these features are lacking context that could contribute to our understanding of their potential relationship with depression.

Interestingly, several studies investigated individual differences in age, gender and occupational status, to inform predictions for individuals. Understanding the impact these differences have on smartphone-related behaviours may enable the development of more personalised digital phenotyping or prevention tools, for example through stratifying individuals into informative groups for detecting recurrence in depression. It is likely that the smartphone-derived features themselves may vary in usefulness between individuals (e.g., some individuals may never use the basic call function, whereas others may regularly make calls using this function). As such, feature selection could perhaps be carried out for each individual for use in individual models, or for subsets of individuals, although it would need to be investigated whether this extra computational step would lead to increases in model performance. Studies were yet to consider other factors that could affect smartphone measures that may impact individual predictions, such as family- or work-related smartphone usage or locations. For example, in the case of an individual who works from home, minimal mobility away from the home location and large call volume are likely unrelated, or rather inseparable, from their mood status. Measures related to expected smartphone usage and lifestyle may, therefore, help to inform smartphone-derived predictions on the individual level and interactions with changing contexts, and could be collected in future studies.

With all of the smartphone-derived features used, it should be noted that digital phenotyping is limited to the assessment of data on behaviours, activities or physical responses that can be passively registered by a smartphone. That is, it cannot assess underlying motives or experiences behind these behaviours, activities or physical responses (unless EMA or self-report are additionally administered). For example, for an individual the qualitative inference can be made that using their smartphone at night may lead to depression, but the more direct relationship could be that the smartphone use during the night might indicate insomnia associated with depression. Without assessments of underlying motives or experiences, digital phenotyping results must be interpreted with care.

### Important study differences

The aim of this review was not to compare smartphone-derived digital phenotypes to other phenotypes, such as wearable-derived digital phenotypes, however as some studies combined smartphone features with other features, we can make some preliminary comments on the effect of combining different types of phenotypes. For example, Pedrelli, et al. (2020) included wearable features, and found that it was inconclusive which modality performed better to predict residual depressive symptom scores. In their models classifying mood states, Bai, et al. (2021) found that their best performing model combined a smartphone feature (call logs) with wearable features (sleep, step count, heart rate), therefore outperforming models using smartphone data alone. To predict treatment response, Zou, et al. (2023) also achieved the best performance in a model using a combination of smartphone and wearable features. Future reviews could seek to compare studies focusing on wearables more generally in predictions for MDD.

Aside from using passive features from other devices, Pellegrini, et al. (2022) chose to use previous depressive symptom scores to predict the following score, finding that this improved prediction. This same study found that including smartphone data in their various models did not always improve predictions, but noted that due to the convenience of smartphone data, smartphone-based models may still be worthwhile (also noted by Cao, et al. (2020)). Kathan, et al. (2022) also found that including active data, in this case EMA data, improved prediction of depressive symptom severity. Zou, et al. (2023) also included baseline and follow-up depressive symptom scores as features in their models. Another study included historical weather data in their model (Pedrelli, et al., 2020), showing that it is possible to include broader contextual information that may affect an individual’s behaviour in models. Interestingly, Pedrelli, et al. (2020) found that individual median HDRS scores provided better predictions than their machine learning models using passive data. Digital phenotyping researchers could seek to incorporate more contextual information in prediction models, including seasonal/time-related information.

Regarding studies that focused on predicting depressive symptom severity, several studies (Braund, et al., 2022; Cao, et al., 2020; Kathan, et al., 2022; Zhang, et al., 2021; Zhang, et al., 2022) used participant-rated depressive symptoms in the form of PHQ scores. Others focused on clinician-rated depressive symptoms, such as the MADRS (Sverdlov, et al., 2021; Pellegrini, et al., 2022) and the HDRS (Pedrelli, et al., 2020). Using participant-rated symptom severity scores may allow for more frequent symptom assessment and therefore a higher temporal resolution of symptom course to be predicted, whereas clinician-rated symptom severity may allow for more consistent measurements between participants, but is less convenient for frequent assessments.

Varying quantities of data (i.e., units of analysis) were defined as a sample by each study, with some studies treating each day as a separate sample. As the commonly used PHQ tool assesses symptoms over a period of 2 weeks, most studies using this tool selected passive smartphone features calculated from the two weeks preceding the administration of the questionnaire. Other studies chose to focus on a participant’s entire data, which allows for longer periods to be analysed. Thus, the unit of analysis that is selected highly depends on research aims or the intended clinical application.

As is the case for other health applications, an open question in digital phenotyping research is whether to develop individual or group models, or perhaps combinations of the two. This review identified studies that predominantly applied group models, with individual factors commonly addressed in models as covariates (Faurholt-Jepsen, et al., 2022; Fujino, Tokuda, & Fujimoto, 2023; Zhang, et al., 2021; Zhang, et al., 2022), or as predictors of interest themselves (Kim, et al., 2023; Laiou, et al., 2022; Pellegrini, et al., 2022). Three studies compared group models to individual models (Cho, et al., 2019; Kathan, et al., 2022; Pedrelli, et al., 2020). In these studies, it was found that individual models often outperformed group models. In terms of developing models that are useful in practice, it may be worthwhile to investigate how other patients’ data may be useful for individual predictions, as varying amounts of data can be expected per patient; for example, some patients may have more than a year’s worth of data available, whereas new patients may only have a few weeks of data available but could still be experiencing clinically significant changes. Models with some shared group parameters (such as in Kathan, et al. (2022)) may therefore be able to contribute to clinical predictions, which could also be informed by appropriate clustering of patients using factors such as employment status and other important individual differences.

### Limitations

This systematic review identified several studies that have made important progress in linking behavioural phenotypes to clinically-relevant variables such as symptom severity and mood state, despite challenges arising from the nature of digital phenotyping data. These included frequent issues with missing data, and the need to combine various high temporal resolution channels in a meaningful way. This often led to the exclusion of large volumes of data and the common use of time-averaged features, risking the loss of useful information relating to temporal dynamics. The digital phenotyping field still needs to achieve higher model performance before the models can be clinically useful without adding additional burden to clinicians in the form of difficult-to-interpret-models or models with low predictive power and consequently high rates of false positive and false negative predictions.

Our review has some limitations, especially with regards to the effect of our search criteria. Studies were restricted to those with MDD populations to avoid too much heterogeneity between different psychiatric populations and/or too general populations with relatively low symptom scores. This may have introduced a selection bias in favour of studies from research groups in WEIRD (‘Western, Educated, Industrialized, Rich, and Democratic’) countries or regions, which could have more resources contributing to their mental health care systems and easier access to a population with diagnosed major depression. This criterion also led to online studies being excluded, as possible MDD diagnosis of participants could not be confirmed using clinical tools. By restricting to studies including an MDD population, this consequently limited the number of studies that could be compared within each analysis goal. In addition, differences in the methodologies used by each of the studies make it more complicated to determine what the overall most predictive variables are for the different goals, limiting the possibilities to make direct comparisons. Future reviews could seek to focus more broadly on studies of depressive symptoms and/or other psychiatric populations within single prediction goals, for a more in-depth comparison of the methods.

### Recommendations

The popularity of digital phenotyping continues to grow as the smartphone maintains its place in today’s world. In order to fully take advantage of digital phenotyping’s clinical potential in MDD, attention should be paid to careful model development.

Firstly, given the changing nature of human behaviour, it is important to acknowledge the temporal dynamics of clinically-relevant changes in the formulation of prediction goals and selection of prediction methods. That is, future approaches may seek to investigate temporal dynamics more directly through choosing models which can handle time series data. Currently only a small number of studies were found to take this approach. Outside of MDD research, a recent paper on predicting schizophrenia relapse using smartphone data applied an anomaly detection approach to investigate whether daily features are anomalous relative to nearby days (Cohen, et al., 2023). To gain further insight into temporal dynamics of individuals’ experiences associated with change, digital phenotyping approaches can be combined with EMA data. A shift towards investigations of temporal dynamics may provide more timely predictions of clinically-relevant changes.

Secondly, before models can reasonably be expected to be used by clinicians in practice, their performance should be improved. Greater collaboration between research groups could allow for larger datasets to be used in model development, and more investigations of generalisability. Replicating results in external validation datasets aids in generalising results to broader settings. As such, increasing efforts to externally validate model performance can help strengthen arguments that digital phenotyping tools can be useful in clinical practice. Kathan et al. (2022) carried out initial investigations of bias in their models; it will be important to ensure models perform as fairly between patients as possible.

Thirdly, models should account for missing data to avoid excessive sample exclusions. To summarise the steps that can be taken to handle the challenge of missing data, efforts can be made to minimise missing data during data collection. Clear instructions should be given to participants so they do not accidentally switch off app functionalities required for collection. However, even if users do not accidentally cause data collection to be impacted, large volumes of missing data can still occur. Incoming data should be regularly inspected to ensure prolonged periods of missing data are not occurring, and researchers can then take action to restore app functionality if this is indeed the case. Data should be inspected across the various sensors that are investigated in case the issue is not affecting all sensors. It could be useful to develop automatic data-checking tools to identify periods of missing data, especially before incorporating prediction models in clinical practice. Even once all has been done to minimise missing data occurring during data collection, there will inevitably still be some instances of missing data that need to be handled. Ideally, minimal participants/samples need to be discarded, although minimum data availability requirements may be needed to filter out participants/samples that are missing large volumes of data. Thresholds for missing value requirements do not necessarily need to be consistent across studies, but could be investigated during model selection/training. For the remaining participants/samples, an appropriate imputation method could be considered (and models with and without imputation compared). Models could also be chosen based on their ability to manage missing data, for example, Hidden Markov Models can accommodate for missing timepoints. Whilst there is not necessarily a one-size-fits-all approach to handling missing data, overall, it seems that to model digital phenotyping data, minimum data availability requirements are needed, and ideally models allowing for data to be missing should be used. The potential bias arising due to the non-randomness of missing data and the optimal strategies used to handle missing data remain unexplored.

## Conclusion

As the field of digital phenotyping develops, we get closer towards the goal of making insightful clinical predictions that can help people with depression, through earlier identification of changes in symptom course and possible onset of future episodes. The studies identified in this review demonstrated moderate success across various prediction goals, including predicting symptom severity and mood state, despite challenges from complex, high-dimensional time series and a high propensity for missing data. Once models with current prediction goals can achieve higher performance across different settings and MDD populations, digital phenotyping research could start to shift towards investigating how to implement these models in practice, for example whether rolling windows should be used to analyse the incoming temporal data. With careful model decisions and implementations, including clinically-and technically-informed feature construction and appropriate validations, digital phenotyping methods for MDD could be generalised to other disorders, with the eventual goal to one day be able to make online predictions of mental disorders that can be directly used by clinicians for improved individualised interventions and patient outcomes.

## Data Availability

This study is a systematic review; all data evaluated in the review are available in the referenced studies. No meta-analysis was carried out.

## References

Abbas, A., Sauder, C., Yadav, V., Koesmahargyo, V., Aghjayan, A., Marecki, S., … Galatzer-Levy, I. R. (2021). Remote digital measurement of facial and vocal markers of major depressive disorder severity and treatment response: a pilot study. Frontiers in digital health, 3, 610006. doi:10.3389/fdgth.2021.610006

Aschbrenner, K. A., Naslund, J. A., Tomlinson, E. F., Kinney, A., Pratt, S. I., & Brunette, M. F. (2019). Adolescents’ use of digital technologies and preferences for mobile health coaching in public mental health settings. Frontiers in public health, 7, 178. doi:10.3389/fpubh.2019.00178

Bai, R., Xiao, L., Guo, Y., Zhu, X., Li, N., Wang, Y., … Wang, G. (2021). Tracking and monitoring mood stability of patients with major depressive disorder by machine learning models using passive digital data: Prospective naturalistic multicenter study. JMIR mHealth and uHealth, 9(3). doi:10.2196/24365

Benoit, J., Onyeaka, H., Keshavan, M., & Torous, J. (2020). Systematic review of digital phenotyping and machine learning in psychosis spectrum illnesses. Harvard Review of Psychiatry, 28, 296–304. doi:10.1097/HRP.0000000000000268

Braund, T. A., Zin, M. T., Boonstra, T. W., Wong, Q. J., Larsen, M. E., Christensen, H., … O’Dea, B. (2022). Smartphone Sensor Data for Identifying and Monitoring Symptoms of Mood Disorders: A Longitudinal Observational Study. JMIR Mental Health, 9(5). doi:10.2196/35549

Buckman, J. E., Underwood, A., Clarke, K., Saunders, R., Hollon, S. D., Fearon, P., & Pilling, S. (2018). Risk factors for relapse and recurrence of depression in adults and how they operate: A four-phase systematic review and meta-synthesis. Clinical psychology review, 64, 13–38. doi:10.1016/j.cpr.2018.07.005

Burcusa, S. L., & Iacono, W. G. (2007). Risk for recurrence in depression. Clinical psychology review, 27, 959–985. 10.1016/j.cpr.2007.02.005

Cao, J., Truong, A. L., Banu, S., Shah, A. A., Sabharwal, A., & Moukaddam, N. (2020). Tracking and predicting depressive symptoms of adolescents using smartphone-based self-reports, parental evaluations, and passive phone sensor data: Development and usability study. JMIR Mental Health, 7(1). doi:10.2196/14045

Cho, C. H., Lee, T., Kim, M. G., In, H. P., Kim, L., & Lee, H. J. (2019). Mood prediction of patients with mood disorders by machine learning using passive digital phenotypes based on the circadian rhythm: Prospective observational cohort study. Journal of Medical Internet Research, 21(4). doi:10.2196/11029

Cohen, A., Naslund, J. A., Chang, S., Nagendra, S., Bhan, A., Rozatkar, A., … others. (2023). Relapse prediction in schizophrenia with smartphone digital phenotyping during COVID-19: a prospective, three-site, two-country, longitudinal study. Schizophrenia, 9, 6. doi:10.1038/s41537-023-00332-5

Emden, D., Goltermann, J., Dannlowski, U., Hahn, T., & Opel, N. (2021). Technical feasibility and adherence of the Remote Monitoring Application in Psychiatry (ReMAP) for the assessment of affective symptoms. Journal of Affective Disorders, 294, 652–660. doi:10.1016/j.jad.2021.07.030

Farhan, A. A., Yue, C., Morillo, R., Ware, S., Lu, J., Bi, J., … Wang, B. (2016). Behavior vs. introspection: refining prediction of clinical depression via smartphone sensing data. 2016 IEEE wireless health (WH), (pp. 1–8). 10.1109/WH.2016.7764553

Faurholt-Jepsen, M., Busk, J., Rohani, D. A., Frost, M., Tønning, M. L., Bardram, J. E., & Kessing, L. V. (2022). Differences in mobility patterns according to machine learning models in patients with bipolar disorder and patients with unipolar disorder. Journal of Affective Disorders, 306, 246–253. doi:10.1016/j.jad.2022.03.054

Fujino, Y., Tokuda, F., & Fujimoto, S. (2023). Decreased step count prior to the first visit for MDD treatment: a retrospective, observational, longitudinal cohort study of continuously measured walking activity obtained from smartphones. Frontiers in public health, 11, 1190464. doi:10.3389/fpubh.2023.1190464

Harari, G. M., Müller, S. R., Aung, M. S., & Rentfrow, P. J. (2017). Smartphone sensing methods for studying behavior in everyday life. Current opinion in behavioral sciences, 18, 83–90. doi:10.1016/j.cobeha.2017.07.018

Higgins, J. P. (2016). A revised tool for assessing risk of bias in randomized trials. Cochrane database of systematic reviews, 10. 10.1002/14651858.CD201601

Kathan, A., Harrer, M., Küster, L., Triantafyllopoulos, A., He, X., Milling, M., … Schuller, B. W. (2022). Personalised depression forecasting using mobile sensor data and ecological momentary assessment. Frontiers in digital health, 4, 964582. doi:10.3389/fdgth.2022.964582

Kim, J. S., Wang, B., Kim, M., Lee, J., Kim, H., Roh, D., … Ryan, N. (2023). Prediction of Diagnosis and Treatment Response in Adolescents With Depression by Using a Smartphone App and Deep Learning Approaches: Usability Study. JMIR formative research, 7, e45991. doi:10.2196/45991

Knights, J., Shen, J., Mysliwiec, V., & DuBois, H. (2023). Associations of smartphone usage patterns with sleep and mental health symptoms in a clinical cohort receiving virtual behavioral medicine care: a retrospective study. Sleep advances : a journal of the Sleep Research Society, 4, zpad027. doi:10.1093/sleepadvances/zpad027

Laiou, P., Kaliukhovich, D. A., Folarin, A. A., Ranjan, Y., Rashid, Z., Conde, P., … Hotopf, M. (2022). The Association between Home Stay and Symptom Severity in Major Depressive Disorder: Preliminary Findings from a Multicenter Observational Study Using Geolocation Data from Smartphones. JMIR mHealth and uHealth, 10(1). doi:10.2196/28095

Lee, H. J., Cho, C. H., Lee, T., Jeong, J., Yeom, J. W., Kim, S., … Kim, L. (2023). Prediction of impending mood episode recurrence using real-time digital phenotypes in major depression and bipolar disorders in South Korea: a prospective nationwide cohort study. Psychological medicine, 53, 5636–5644. doi:10.1017/S0033291722002847

Lim, S. S., Vos, T., Flaxman, A. D., Danaei, G., Shibuya, K., Adair-Rohani, H., … others. (2012). A comparative risk assessment of burden of disease and injury attributable to 67 risk factors and risk factor clusters in 21 regions, 1990–2010: a systematic analysis for the Global Burden of Disease Study 2010. The lancet, 380, 2224–2260. doi:10.1016/S0140-6736(12)61766-8

Luo, W., Phung, D., Tran, T., Gupta, S., Rana, S., Karmakar, C., … others. (2016). Guidelines for developing and reporting machine learning predictive models in biomedical research: a multidisciplinary view. Journal of medical Internet research, 18, e323. doi:10.2196/jmir.5870

Maher, N. A., Senders, J. T., Hulsbergen, A. F., Lamba, N., Parker, M., Onnela, J.-P., … Broekman, M. L. (2019). Passive data collection and use in healthcare: A systematic review of ethical issues. International journal of medical informatics, 129, 242–247. 10.1016/j.ijmedinf.2019.06.015

Matcham, F., Leightley, D., Siddi, S., Lamers, F., White, K. M., Annas, P., … Hotopf, M. (2022). Remote Assessment of Disease and Relapse in Major Depressive Disorder (RADAR-MDD): recruitment, retention, and data availability in a longitudinal remote measurement study. BMC Psychiatry, 22(1). doi:10.1186/s12888-022-03753-1

Müller, S. R., Chen, X., Peters, H., Chaintreau, A., & Matz, S. C. (2021). Depression predictions from GPS-based mobility do not generalize well to large demographically heterogeneous samples. Scientific Reports, 11, 14007. doi:10.1038/s41598-021-93087-x

Nelson, B. W., & Allen, N. B. (2018). Extending the passive-sensing toolbox: using smart-home technology in psychological science. Perspectives on psychological science, 13, 718–733. doi:10.1177/1745691618776008

Pedrelli, P., Fedor, S., Ghandeharioun, A., Howe, E., Ionescu, D. F., Bhathena, D., … Picard, R. W. (2020). Monitoring Changes in Depression Severity Using Wearable and Mobile Sensors. Frontiers in Psychiatry, 11. doi:10.3389/fpsyt.2020.584711

Pellegrini, A. M., Huang, E. J., Staples, P. C., Hart, K. L., Lorme, J. M., Brown, H. E., … Onnela, J. P. (2022). Estimating longitudinal depressive symptoms from smartphone data in a transdiagnostic cohort. Brain and Behavior, 12(2). doi:10.1002/brb3.2077

Saeb, S., Zhang, M., Karr, C. J., Schueller, S. M., Corden, M. E., Kording, K. P., … others. (2015). Mobile phone sensor correlates of depressive symptom severity in daily-life behavior: an exploratory study. Journal of medical Internet research, 17, e4273. doi:10.2196/jmir.4273

Siddi, S., Giné-Vázquez, I., Bailon, R., Matcham, F., Lamers, F., Kontaxis, S., … Consortium, O. B.-C. (2022). Biopsychosocial Response to the COVID-19 Lockdown in People with Major Depressive Disorder and Multiple Sclerosis. Journal of clinical medicine, 11. doi:10.3390/jcm11237163

Silva, W. J., Lopes, L., Galdino, M. K., & Almeida, A. A. (2021). Voice acoustic parameters as predictors of depression. Journal of Voice. 10.1016/j.jvoice.2021.06.018

Sun, S., Folarin, A. A., Zhang, Y., Cummins, N., Garcia-Dias, R., Stewart, C., … Dobson, R. J. (2023). Challenges in Using mHealth Data From Smartphones and Wearable Devices to Predict Depression Symptom Severity: Retrospective Analysis. Journal of medical Internet research, 25, e45233. doi:10.2196/45233

Sverdlov, O., Curcic, J., Hannesdottir, K., Gou, L., Luca, V. D., Ambrosetti, F., … Jacobs, G. E. (2021). A Study of Novel Exploratory Tools, Digital Technologies, and Central Nervous System Biomarkers to Characterize Unipolar Depression. Frontiers in Psychiatry, 12. doi:10.3389/fpsyt.2021.640741

Tønning, M. L., Faurholt-Jepsen, M., Frost, M., Bardram, J. E., & Kessing, L. V. (2021). Mood and Activity Measured Using Smartphones in Unipolar Depressive Disorder. Frontiers in Psychiatry, 12. doi:10.3389/fpsyt.2021.701360

Ware, S., Yue, C., Morillo, R., Lu, J., Shang, C., Bi, J., … Wang, B. (2020). Predicting depressive symptoms using smartphone data. Smart Health, 15, 100093. doi:10.1016/j.smhl.2019.100093

Wasserzug, Y., Degani, Y., Bar-Shaked, M., Binyamin, M., Klein, A., Hershko, S., & Levkovitch, Y. (2023). Development and validation of a machine learning-based vocal predictive model for major depressive disorder. Journal of affective disorders, 325, 627–632. doi:10.1016/j.jad.2022.12.117

Zhang, Y., Folarin, A. A., Sun, S., Cummins, N., Ranjan, Y., Rashid, Z., … Dobson, R. J. (2021). Predicting depressive symptom severity through individuals’ nearby bluetooth device count data collected by mobile phones: Preliminary longitudinal study. JMIR mHealth and uHealth, 9(7). doi:10.2196/29840

Zhang, Y., Folarin, A. A., Sun, S., Cummins, N., Vairavan, S., Bendayan, R., … Dobson, R. J. (2022). Longitudinal Relationships Between Depressive Symptom Severity and Phone-Measured Mobility: Dynamic Structural Equation Modeling Study. JMIR Mental Health, 9(3). doi:10.2196/34898

Zhang, Y., Pratap, A., Folarin, A. A., Sun, S., Cummins, N., Matcham, F., … Dobson, R. J. (2023). Long-term participant retention and engagement patterns in an app and wearable-based multinational remote digital depression study. NPJ digital medicine, 6, 25. doi:10.1038/s41746-023-00749-3

Zou, B., Zhang, X., Xiao, L., Bai, R., Li, X., Liang, H., … Wang, G. (2023). Sequence Modeling of Passive Sensing Data for Treatment Response Prediction in Major Depressive Disorder. IEEE transactions on neural systems and rehabilitation engineering : a publication of the IEEE Engineering in Medicine and Biology Society, 31, 1786–1795. doi:10.1109/TNSRE.2023.3260301

