## Supplementary material for "From smartphone data to clinically relevant predictions: A systematic review of digital phenotyping methods in depression"

**Supplementary Materials**

**S1. Key words and search strings per database**

Search strings for the various electronic databases (i.e., PubMed, Embase, and PsycINFO) were investigated to establish reasonable searches. These searches contain three main elements:

The search is restricted to studies that focus on MDD, e.g.:

"major depressive disorder"

"MDD"

"depression"

And that are related to digital phenotyping or monitoring, e.g.:

"digital phenotyp*"

"monitor*"

"sensor"

"track*"

And finally, restricted to studies related to (smart)phones through searching for:

“phone*”

“smartphone*”

“mobile*”

For all databases, the searches were restricted to studies published from 2012-November 2023. No filters were used. Complete search strings with keywords for each database are specified below.

**PubMed**

Block 1: (("Depression"[Mesh] OR "Depressive Disorder, Major"[Mesh] OR MDD[tiab] OR depression[tiab] OR depressive[tiab])

AND

Block 2: ("Cell Phone"[Mesh] OR "Computers, handheld"[Mesh] OR "Telemedicine"[Mesh] OR android*[tiab] OR app[tiab] OR apps[tiab] OR cellphone*[tiab] OR ios[tiab] OR iphone*[tiab] OR mobile*[tiab] OR phone*[tiab] OR smartphone*[tiab])

AND

Block 3: ("Monitoring, Ambulatory"[Mesh] OR digital phenotyp*[tiab] OR monitor*[tiab] OR sensing[tiab] OR sensor*[tiab] OR track*[tiab]))

AND 2012/01/01:{current date}[Date - Publication]

**PsycINFO**

Block 1: ((exp major depression/ OR MDD.ti,ab,id. OR depression.ti,ab,id. OR depressive.ti,ab,id.)

AND

Block 2: (exp mobile devices/ OR exp mobile health/ OR exp mobile technology/ OR android*.ti,ab,id. OR app.ti,ab,id. OR apps.ti,ab,id. OR cellphone*.ti,ab,id. OR ios.ti,ab,id. OR iphone*.ti,ab,id. OR mobile*.ti,ab,id. OR phone*.ti,ab,id. OR smartphone*.ti,ab,id.)

AND

Block 3: (exp activity level/ OR exp monitoring/ OR exp tracking/ OR "digital phenotyp*".ti,ab,id. OR monitor*.ti,ab,id. OR sensing.ti,ab,id. OR sensor*.ti,ab,id. OR track*.ti,ab,id.))

Publication year = 2012 – Current

**Embase**

Block 1: ((exp depression/ OR MDD. ti,ab,kf. OR depression. ti,ab,kf. OR depressive. ti,ab,kf.)

AND

Block 2: (exp "cell phone use"/ OR exp mobile application/ OR exp mobile phone/ OR exp telemedicine/ OR android*.ti,ab,kf. OR app.ti,ab,kf. OR apps.ti,ab,kf. OR cellphone*.ti,ab,kf. OR ios.ti,ab,kf. OR iphone*.ti,ab,kf. OR mobile*.ti,ab,kf. OR phone*.ti,ab,kf. OR smartphone*.ti,ab,kf.)

AND

Block 3: (exp ambulatory monitoring/ OR "digital phenotyp*".ti,ab,kf. OR monitor*.ti,ab,kf. OR sensing.ti,ab,kf. OR sensor*.ti,ab,kf. OR track*.ti,ab,kf.))

Publication year = 2012 - Current

**Scopus**

Block 1: ((TITLE-ABS-KEY(depression OR depressive OR MDD))

AND

Block 2: (TITLE-ABS-KEY(android* OR app OR apps OR cellphone* OR ios OR iphone* OR mobile* OR phone* OR smartphone*))

AND

Block 3: (TITLE-ABS-KEY("digital phenotyp*" OR monitor* OR sensing OR sensor* OR track*)))

AND PUBYEAR > 2011

AND ( LIMIT-TO ( DOCTYPE , "ar" ) )

**Web of Science**

Block 1: (TS=(depression OR depressive OR MDD)

AND

Block 2: TS=(android* OR app OR apps OR cellphone* OR ios OR iphone* OR mobile* OR phone* OR smartphone*)

AND

Block 3: TS=("digital phenotyp*" OR monitor* OR sensing OR sensor* OR track*))

Timespan: 2012-01-01 to {current date} (Publication Date)

Document Types: Articles

**S2. Data extraction form**

Study characteristics:

•	Author

•	Year of publication

•	Country

•	Population

•	% Female

•	Age (Mean)

•	Ethnicity

•	Sample size

General methodological information:

•	Expected participation duration

•	What did a single measurement refer to?

•	Outlier removal

•	Handling of missing values

•	Specific variable/feature processing, selection, generation, or reduction methods used

Specific methodological details:

•	Goal of analysis

•	Types of predictor variables

•	Response variable/s

•	Modelling techniques

•	Quality metrics used to assess model performance

•	(Internal) validation/assessment strategy

•	(Internal) validation/assessment metrics

•	Results

**S3. Risk of Bias Assessment**

**Risk of bias rating per study: Subdomains rating using the Cochrane Collaboration Risk of Bias (RoB)**

| Author (year of publication) | Domain 1  *Random sequence generation* | Domain 2  *Allocation concealment* | Domain 3  *Blinding of participants/ personnel* | Domain 4  *Blinding of outcome assessment* | Domain 5  *Incomplete outcome data* | Domain 6  *Selective reporting* |
| --- | --- | --- | --- | --- | --- | --- |
| Bai et al. (2021) | NA | NA | NA | NA | Some concerns | Low risk |
| Braund et al. (2022) | NA | NA | NA | NA | Some concerns | Low risk |
| Cao et al. (2020) | NA | NA | NA | NA | Some concerns | Low risk |
| Cho et al. (2019) | NA | NA | NA | NA | High risk | Low risk |
| Emden et al. (2020) | NA | NA | NA | NA | NA | Low risk |
| Faurholt-Jepsen et al. (2022) | Low risk | Low risk | NA | Some concerns | High risk | Low risk |
| Fujino et al. (2023) | NA | NA | NA | NA | Some concerns | Low risk |
| Kathan et al. (2022) | NA | NA | NA | NA | High risk | High risk |
| Kim et al. (2023) | NA | NA | NA | NA | High risk | Low risk |
| Knights et al. (2023) | NA | NA | NA | NA | Low risk | Low risk |
| Laiou et al. (2021) | NA | NA | NA | NA | Some concerns | Low risk |
| Lee et al. (2023) | NA | NA | NA | NA | Some concerns | Low risk |
| Matcham et al. (2022) | NA | NA | NA | NA | NA | Low risk |
| Pedrelli et al. (2020) | NA | NA | NA | NA | Some concerns | Low risk |
| Pellegrini et al. (2021) | NA | NA | NA | NA | Some concerns | Some concerns |
| Siddi et al. (2022) | NA | NA | NA | NA | Some concerns | Low risk |
| Sun et al. (2023) | NA | NA | NA | NA | Low risk | Low risk |
| Sverdlov et al. (2021) | NA | NA | NA | NA | High risk | Low risk |
| Tønning et al. (2021) | Low risk | Low risk | NA | Some concerns | Some concerns | Low risk |
| Wasserzug et al. (2023) | NA | NA | NA | NA | Low risk | Low risk |
| Zhang et al. (2021) | NA | NA | NA | NA | Some concerns | Low risk |
| Zhang et al. (2022) | NA | NA | NA | NA | Some concerns | Low risk |
| Zhang et al. (2023) | NA | NA | NA | NA | NA | Low risk |
| Zou et al. (2023) | NA | NA | NA | NA | High risk | Low risk |

**S4. Quality Assessment**

**Quality Assessment per study: Item rating based on** **Luo, et al. (2016), Benoit, Onyeaka, Keshavan, and Torous (2020)**

| Author (year of publication) | Domain 1  *Strategy for handling outliers described* | Domain 2  *Strategy for handling missing values described* | Domain 3  *Summary statistics reported* | Domain 4  *Internal validation strategy described* | Domain 5  *External validation strategy described* |
| --- | --- | --- | --- | --- | --- |
| Bai et al. (2021) | No | Yes | Yes | Yes | NA |
| Braund et al. (2022) | No | No | Yes | NA | NA |
| Cao et al. (2020) | No | No | Yes | NA | NA |
| Cho et al., (2019) | No | Yes | Yes | Yes | NA |
| Emden et al. (2020) | NA | Yes | Yes | NA | NA |
| Faurholt-Jepsen et al. (2022) | Yes | Yes | Yes | Yes | NA |
| Fujino et al. (2023) | No | Yes | Yes | NA | NA |
| Kathan et al. (2022) | Yes | Yes | Yes | Yes | NA |
| Kim et al. (2023) | No | No | Yes | Yes | NA |
| Knights et al. (2023) | No | Yes | Yes | NA | NA |
| Laiou et al. (2021) | Yes | Yes | Yes | Yes | NA |
| Lee et al. (2023) | No | Yes | Yes | Yes | NA |
| Matcham et al. (2022) | NA | Yes | Yes | NA | NA |
| Pedrelli et al. (2020) | No | Yes | Yes | Yes | NA |
| Pellegrini et al. (2021) | No | Yes | Yes | Yes | NA |
| Siddi et al. (2022) | No | Yes | Yes | NA | NA |
| Sun et al. (2023) | Yes | Yes | Yes | NA | NA |
| Sverdlov et al. (2021) | No | Yes | Yes | Yes | NA |
| Tønning et al. (2021) | No | Yes | Yes | NA | NA |
| Wasserzug et al. (2023) | Yes | No | Yes | Yes | NA |
| Zhang et al. (2021) | No | Yes | Yes | Yes | NA |
| Zhang et al. (2022) | Yes | Yes | Yes | NA | NA |
| Zhang et al. (2023) | No | Yes | Yes | NA | NA |
| Zou et al. (2023) | No | Yes | Yes | Yes | NA |

**S5. Ranges in scores for symptom scales**

**Possible ranges in scores for the various symptom scales**

| Symptom scale | Possible range in values |
| --- | --- |
| MADRS | 0-60 |
| HDRS | 0-52 |
| PHQ-9 | 0-27 |
| PHQ-8 | 0-24 |
| PHQ-2 | 0-6 |
